# Enhanced expression of HLA-DR and CD69 on peripheral CD4^+^ T cells predicts better clinical outcomes in cutaneous melanoma

**DOI:** 10.64898/2026.03.24.26349163

**Authors:** Ana Tomás, José Maximino, Hugo Nunes, Rute Salvador, Rafael Luís, Cheila Brito, Diana P. Saraiva, Emanuel Gouveia, Carolina Pereira, Filipe Gonçalves, Victor Farricha, Elisabete Lopez Carvalho, Cecília Moura, Maria José Passos, Sofia Cristóvão-Ferreira, Patrícia M. Pereira, Maria Guadalupe Cabral, Marta Pojo

## Abstract

**Background:** Cutaneous melanoma (CM) is an aggressive skin cancer with rising incidence, representing a growing public health concern. Despite the remarkable success of immune-checkpoint inhibitors (ICIs) in the management of advanced disease, mortality remains high due to therapy resistance. Identifying reliable prognostic and predictive biomarkers is therefore essential to improve patient stratification, optimize treatment selection, and minimize unnecessary toxicity.

**Methods:** We comprehensively profiled the circulating immune landscape of 54 treatment-naïve CM patients by integrating flow cytometry immunophenotyping with clinicopathological data, and performed tumor gene expression analysis in a subset of 26 patients.

**Results:** Elevated HLA-DR and CD69 expression on circulating CD4^+^ T cells, together with reduced circulating CD8^+^ T cell frequency, emerged as candidate prognostic biomarkers associated with improved survival. Prognostic models combining these immune variables with clinical covariates accurately stratified patients by overall survival (89.5% sensitivity, 72.7% specificity; AUC = 0.872, *p* < 0.0001) and progression/recurrence risk (75% sensitivity and 71.4% specificity; AUC = 0.763, *p* = 0.001). In a subset of 43 patients subsequently treated with ICIs, elevated baseline HLA-DR and CD69 expression on circulating CD4^+^ T cells was also associated with therapeutic benefit. A predictive model integrating these markers with clinical covariates achieved good discriminatory performance (65.2% sensitivity, 88.9% specificity; AUC = 0.775, *p* = 0.0027). Tumor gene expression profiling supported the role of IFN-γ-related signatures, previously linked to ICI response, as complementary prognostic and predictive tools.

**Conclusion:** These findings highlight systemic CD4^+^ T cell activation status as a promising, easily measurable biomarker in CM, laying the foundation for future strategies to refine patient stratification and guiding immunotherapy decisions.

## 1. Introduction

Cutaneous melanoma (CM) is the deadliest skin cancer, accounting for over 60% of skin cancer-related deaths despite representing less than 5% of cases [1–3]. Nearly 60,000 deaths were reported worldwide in 2022, and incidence continues to rise, with projections of 500,000 new cases annually by 2040 [2, 4]. While early-stage disease (I−IIA) is often curable by surgery alone, stages IIB−IV typically require systemic therapy [5].

Over the past decade, therapeutic advances have significantly improved CM outcomes [6]. Targeted therapy with BRAF and MEK inhibitors achieves high initial response rates in *BRAF*-mutant tumors, but resistance limits long-term benefit [6, 7]. Immune-checkpoint inhibitors (ICIs) targeting PD-1 (nivolumab, pembrolizumab) and CTLA-4 (ipilimumab) have significantly improved survival, with combination regimens achieving 10-year overall survival (OS) up to 43% in unresectable CM [8]. ICIs are now standard first-line therapy for unresectable CM and are also approved as adjuvant treatment for resected high-risk stage IIB/C−III disease [5, 9].

Despite these advances, nearly half of patients fail to respond to ICIs, and many experience immune-related toxicity [8, 10]. Reliable biomarkers are therefore needed to guide treatment selection, minimize toxicity, and reduce costs. In metastatic CM, predictive markers could identify patients unlikely to benefit from ICIs, enabling earlier consideration of alternative strategies. Current research primarily focuses on tumor-derived predictive markers, including tumour mutational burden [11–13], PD-L1 expression [14–16], tumor-infiltrating lymphocytes [17–19], and IFN-γ-related gene signatures [12, 20–22]. Yet, none have demonstrated consistent clinical utility [5, 23–28]. Likewise, prognostic biomarkers remain inadequate, as outcomes vary widely among patients within the same stage [29]. Robust prognostic markers could identify patients requiring adjuvant therapy or closer surveillance, while sparing low-risk individuals from overtreatment.

Most biomarker approaches rely on tumor tissue, which is limited by heterogeneity and restricted accessibility [30, 31]. Peripheral blood represents a minimally invasive, reproducible, and dynamic alternative. However, serum lactate dehydrogenase (LDH) and S100B remain the only circulating biomarkers in clinical use, and their prognostic value is limited [32–34]. Identifying reliable blood-based biomarkers remains a critical unmet need in CM.

Tumor progression and ICI response depend on coordinated interactions between tumor and immune cells, with systemic immunity increasingly recognized as a key determinant of immunotherapy efficacy [35]. Circulating immune profiles may reflect the tumor microenvironment (TME), yet most prior studies in CM have been limited by small cohorts (15−40 patients), a focus on metastatic disease and ICI response, and an emphasis on CD8^+^ T cells [36–43]. Moreover, immunosuppressive features such as regulatory T cells or inhibitory molecules have been largely overlooked.

To address these gaps, we performed comprehensive profiling of circulating immune cell subsets in treatment-naïve CM patients, integrated with clinicopathological data and longitudinal follow-up. In a subset of patients subsequently treated with anti-PD-1 therapy, baseline immune features were also evaluated as predictors of clinical benefit, in both palliative and adjuvant settings. Complementary tumor gene expression analysis was also conducted. Our findings reveal circulating CD4^+^ T cells expressing high levels of activation markers HLA-DR and CD69 as candidate prognostic and predictive biomarkers in CM, offering a practical, minimally invasive tool to refine patient stratification and elucidate systemic antitumor immunity.

## 2. Materials and Methods

### 2.1. Patients and study design

A total of 85 CM patients, stages IIC−IV, were prospectively recruited at Instituto Português de Oncologia de Lisboa Francisco Gentil (IPOLFG) between August 2019 and November 2023. Eligibility criteria included histologically confirmed CM, age ≥ 18 years, and no contraindications to immunotherapy. Peripheral blood and/or tumor samples were collected during routine clinical procedures, and anonymized clinicopathological and follow-up data were obtained. Follow-up was completed by June 2025.

Peripheral blood was available from 65 patients, including 54 treatment-naïve at the time of collection (Supplementary Table S1). Among these, 43 later received ICIs, and their pre-treatment samples were defined as baseline for predictive analysis (treatment regimens and response criteria in Supplementary Table S2). Therapeutic benefit was assessed according to treatment setting: in the adjuvant setting, benefit corresponded to disease-free status for ≥ 24 months; in the palliative setting, benefit was defined as complete or partial response, or stable disease lasting ≥ 6 months, whereas progressive disease or stable disease < 6 months was considered no benefit.

Longitudinal peripheral blood samples were obtained from 31 of the 43 ICI-treated patients at approximately 3-month intervals. Six additional patients were sampled during ICI therapy without baseline data, resulting in 49 patients with pre- and/or on-treatment samples.

Tumor samples were collected from 31 CM patients at surgical resection of the primary tumor (n = 2), lymph node (n = 15), cutaneous (n = 12), or soft tissue metastases (n = 2), regardless of prior treatment. Paired blood-tumor samples were available for 11 patients (6 lymph node, 4 cutaneous, 1 soft tissue metastases). An overview of patient distribution, sample types, and downstream analysis is shown in Supplementary Figure S1.

Peripheral blood from 15 healthy donors (8 males, 7 females; median age 47 years) was collected at the IPOLFG Hematology Department in January 2022. Donors under 18 years or with known/suspected disease were excluded.

Clinical endpoints were defined as follows: OS as the time from sample collection to death or last follow-up; event-free survival (EFS) as the time to progression, recurrence, or last follow-up; progression-free survival (PFS) as the time to progression or last follow-up, particularly in stage IV patients, and disease-free survival (DFS) as the time to recurrence or last follow-up, particularly in stage IIC/III patients.

Patients included in this study were recruited under a broader research project approved by the IPOLFG Ethics Board Committee (UIC/1310), which also generated a previously published clinical case report describing the establishment of a novel melanoma cell line [44], and a master’s dissertation investigating *ARL1* expression in tumor samples from this cohort [45]. The present study is the first to comprehensively characterize the immune profile of this cohort and to identify immune-based biomarkers associated with clinical outcomes.

This study was also approved by the NOVA Medical School Ethics Committee (n° 128/2023/CEFCM) and conducted in accordance with the Declaration of Helsinki and Portuguese law. Written informed consent was obtained from all participants.

### 2.2. Sample processing

Peripheral blood was collected into BD Vacutainer® K2EDTA tubes (BD Biosciences, 367863). Plasma was separated by centrifugation (3,000 × *g*, 15 min) and stored at −80 °C. Peripheral blood mononuclear cells (PBMCs) were isolated from 12 mL of blood by density gradient centrifugation using Lymphoprep™ (Serumwerk Bernburg, 1114544). Whole blood was diluted 1:1 in phosphate-buffered saline (PBS 1×), layered onto Lymphoprep™ (1:1), and centrifuged (800 × *g*, 30 min, without brake). PBMCs were washed and red blood cells (RBC) lysed with RBC lysis buffer 1× (BioLegend, 420301) for 15 min at room temperature. After a final wash step, PBMCs were cryopreserved at 2 × 10^6^ cells/mL in fetal bovine serum (FBS, Corning 35-079-CV) with 10% dimethyl sulfoxide (DMSO), and stored at −80 °C or in liquid nitrogen.

Fresh tumor specimens were transported in saline buffer and processed immediately. Tissue was mechanically dissociated into 1−2 mm pieces and divided for RNA extraction and/or flow cytometry. Samples for RNA analysis were stored in RNAlater™ (Qiagen) at −80 °C, and those for flow cytometry analysis were cryopreserved in FBS with 10% DMSO.

### 2.3. Flow cytometry

Single-cell suspensions from tumor tissue were prepared using BD Medicon devices (BD Biosciences, 340591) and filtered through 30 µm CellTrics™ mesh filters (Sysmex, 04-004-2326). Thawed PBMCs (4 × 10^6^ cells) or tumor suspensions were stained with Zombie Aqua™ Fixable Viability Dye (BioLegend, 423102; 1:200) in PBS 1× for 30 min, washed in PBS with 2% FBS and 1 mM EDTA, and incubated with surface antibodies (1:100 in PBS 1×; Supplementary Table S3) for 15 min at room temperature. After washing, samples requiring intracellular staining were fixed and permeabilized (eBioscience™ Fixation/Permeabilization Kit, 00-5123-43), incubated with intracellular antibodies (1:100 in Permeabilization Buffer 1×, eBioscience™ 00-8333-56) for 30 min at room temperature, and washed in PBS 1×.

Data were acquired on a BD FACS Canto II using FACSDiva v8.0.1 (BD Biosciences) and analyzed in FlowJo vX.0.7 (BD Biosciences). Samples were processed in randomized batches. Gating strategies are detailed in Supplementary Figure S2. Immune populations are represented as a percentage of live cells and included CD4^+^ T cells (CD3^+^CD4^+^CD8^−^), CD8^+^ T cells (CD3^+^CD8^+^CD4^−^), regulatory T cells (Tregs; CD3^+^CD4^+^CD25^high^CD127^low^), total monocytes (CD45^+^CD68^+^CD14^+^ and/or CD16^+^) and macrophages (CD45^+^CD68^+^). Monocyte subsets were defined as classical (CD45^+^CD68^+^CD14^+^CD16^−^), intermediate (CD45^+^CD68^+^CD14^+^CD16^+^) and non-classical (CD45^+^CD68^+^CD14^−^CD16^+^). Functional marker expression was quantified as the ratio of the median fluorescence intensity (MFI) between positive and negative populations, defined using unstained controls. Subsets were evaluated only when ≥ 300 parent events were available.

### 2.4. RNA extraction and gene expression analysis

RNA was successfully extracted from 26 tumor samples. Tissue was homogenized in TRIzol® reagent (Invitrogen, 15596026), and total RNA isolated following the manufacturer’s protocol, with overnight isopropanol precipitation at 4 °C. Complementary DNA (cDNA) was synthesized from 1 µg of total RNA using SuperScript™ II Reverse Transcriptase (Invitrogen, 18064014) in a 20 µL reaction, then diluted 1:1 with RNase-free water. Reverse transcription quantitative PCR (RT-qPCR) was performed in triplicate using 1 µL of diluted cDNA, 4 µL of SYBR™ Green PCR Master Mix (Applied Biosystems, 4309155), and 0.15 µL of each primer (10 µM), on a LightCycler® 480 System (Roche), as described elsewhere [46]. Primer sequences are listed in Supplementary Table S4. Relative gene expression was calculated using the comparative Ct method (ΔΔCt) [47], with *TBP* as reference.

### 2.5. Cytokine analysis

Plasma IFN-γ levels were measured using the ELISA MAX™ Human IFN-γ Kit (BioLegend, 430101), following manufacturer’s instructions. The capture antibody was diluted 1:200 in sodium carbonate-bicarbonate buffer (100 mM NaHCO_3_, 33.6 mM Na_2_CO_3_, pH 9.5). Washing steps used PBS 0.05% Tween-20, and blocking was performed with PBS 1% bovine serum albumin (BSA). Absorbance was read at 450 nm on an iMark™ Microplate Absorbance Reader (Bio-Rad), and IFN-γ concentrations were interpolated from the standard curve.

### 2.6. Statistical analysis

Statistical analysis was performed in IBM® SPSS® Statistics v25 and GraphPad Prism v9.0.0. Two-tailed tests were used, with *p* < 0.05 considered significant.

Differences between two groups were assessed by the Mann-Whitney U test, and comparisons among multiple groups using the Kruskal-Wallis test followed by Dunn’s correction. Results are shown as median with interquartile ranges. Correlations were assessed using Spearman’s rank test.

Associations between immune variables and clinical outcomes were first tested by univariable Cox proportional hazards (OS, EFS) or logistic regression (therapeutic benefit). Variables with *p* < 0.1 were considered for multivariable analysis. ROC curves assessed the prognostic/predictive performance of selected variables, with optimal cut-offs defined by the maximized Youden’s index. Kaplan-Meier curves were compared using the log-rank test, while Fisher’s exact test determined the groups’ association with therapeutic benefit. Each selected immune variable was entered into multivariable models adjusted for sex, age, stage, and *BRAF* status. Due to missing data, Breslow thickness, ulceration, and histological subtype were excluded as confounders. Patients lacking *BRAF* data (n = 2) were also excluded.

To test combined prognostic/predictive value, a clinical risk score was derived from Cox or logistic models including only confounders. The resulting linear predictor (Xβ) was incorporated with selected immune variables in the final multivariable models. Model performance was evaluated by ROC analysis using linear predictor (prognostic) or predicted probability (predictive). Optimal cut-offs stratified patients for log-rank or Fisher’s exact tests.

For tumor gene expression data, unsupervised hierarchical clustering was performed using Euclidean distance and complete linkage. Associations between clusters and categorical variables were tested using Fisher’s exact test, and survival differences assessed using Kaplan-Meier analysis with log-rank testing. Differential gene expression was analyzed using multiple Mann-Whitney U tests with Benjamini-Krieger-Yekutieli correction.

## 3. Results

### 3.1. Circulating immune profiling reveals functional alterations in advanced CM

Given the immunogenic nature of CM [48], we first assessed whether CM alters circulating immune profiles and whether disease stage further deepens these changes. Major T cell and monocyte subsets were quantified in 15 healthy donors and 53 treatment-naïve CM patients (one stage IIC patient excluded). Overall, subset frequencies were broadly similar between healthy donors and CM patients, with no significant differences between stage III and IV CM (Supplementary Figure S3).

To explore functional alterations, we analyzed activation and polarization markers within each subset (**Figure 1**). Compared with stage III disease, stage IV patients exhibited reduced HLA-DR expression on CD4^+^ T cells and increased CD69 expression on CD8^+^ T cells, both markers of T cell activation [49] (**Figure 1A, H**). Notably, HLA-DR expression, considered a marker of late T cell activation [49], on CD4^+^ T cells was also significantly lower in stage IV patients compared with healthy controls (**Figure 1A**). In contrast, the expression of the remaining T cell markers did not differ significantly between groups (**Figure 1B−G, I, J**). In the monocyte compartment, expression of both CD80 and CD163 was decreased in CM relative to healthy donors (**Figure 1K, L**), while no significant difference was observed for CD206 (**Figure 1M**).

**Figure 1.**
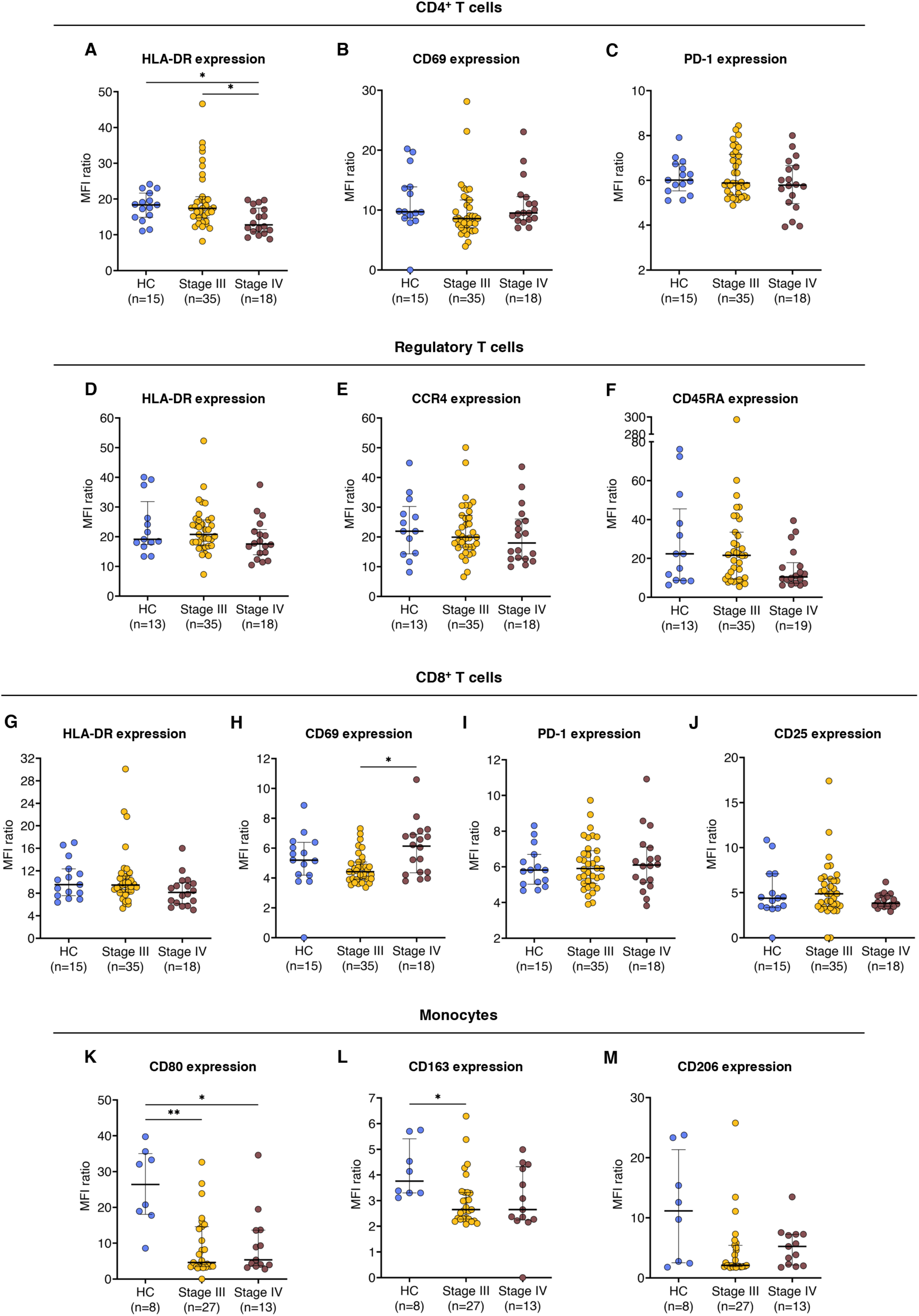
Expression of immunophenotypic markers on circulating immune cell subsets is broadly comparable across cutaneous melanoma stages, with selected functional shifts. Expression of functional markers was quantified by flow cytometry in peripheral blood mononuclear cells (PBMCs) from 15 healthy controls (HC) and 53 cutaneous melanoma (CM) patients (divided into stages III and IV). Expression levels are defined as the median fluorescence intensity (MFI) ratio between marker-positive and -negative populations. Analyzed markers include (A) HLA-DR, (B) CD69, and (C) PD-1 on CD4^+^ T cells; (D) HLA-DR, (E) CCR4, and (F) CD45RA on regulatory T cells (Tregs); (G) HLA-DR, (H) CD69, (I) PD-1, and (J) CD25 on CD8^+^ T cells; (K) CD80, (L) CD163, and (M) CD206 on monocytes. Monocyte subset analysis was limited to 41 patients and 8 healthy controls due to low monocyte frequencies. Each dot represents an individual. Data are shown as median with interquartile range. Statistical comparisons were performed using the Kruskal-Wallis test followed by Dunn’s multiple comparisons test. *, *p* < 0.05; **, *p* < 0.01.

Potential confounders (sex, age, *BRAF* mutational status) were examined (Supplementary Figures S4, S5; Supplementary Table S5), and no major differences related to age or *BRAF* status were observed. Female patients exhibited higher CD4^+^ T cell frequencies, resulting in higher CD3^+^ T cell proportions and increased CD4^+^:CD8^+^ T cell ratio. However, activation marker expression on CD4^+^ T cells did not differ by sex.

Overall, immune subset frequencies were largely comparable between healthy individuals and CM patients and remained stable across disease stages; in contrast, functional alterations, particularly involving T cell activation and monocyte polarization markers, were associated with more advanced CM.

### 3.2. CD4^+^ T cell activation and CD8^+^ T cell frequency in blood as potential prognostic indicators in CM

To identify potential prognostic blood biomarkers, associations between circulating immune variables and clinical outcomes were evaluated in 54 treatment-naïve CM patients using univariable Cox regression (Supplementary Table S6). Among clinicopathological variables, advanced age and stage IV disease were linked to worse OS (HR = 1.037, *p* = 0.025; HR = 2.595, *p* = 0.016). Among immune features, increasing CD8^+^ T cell frequency was significantly associated with poorer OS (HR = 1.049, *p* = 0.010); however, none of the functional markers assessed on CD8^+^ T cells showed potential associations with OS, leaving the functional status of this expanded compartment unclear. Likewise, an increasing CD8^+^:Treg ratio indicated worse outcomes, whereas a higher CD4^+^:CD8^+^ T cell ratio tended to associate with improved OS. Increasing HLA-DR (HR = 0.942, *p* = 0.071) and CD69 (HR = 0.899, *p* = 0.090) expression on CD4^+^ T cells showed a trend toward a protective effect, consistent with reduced HLA-DR levels on CD4^+^ T cells observed in advanced disease. Only HLA-DR on CD4^+^ T cells significantly correlated with longer EFS (HR = 0.913, *p* = 0.015).

Correlation analysis (Supplementary Figure S6) confirmed that HLA-DR and CD69 expression on CD4^+^ T cells were not significantly correlated, reflecting distinct activation states – CD69 is considered an early T cell activation marker, while HLA-DR is typically expressed later [49]. Conversely, CD8^+^ T cell frequency strongly correlated with the CD8^+^:Treg and CD4^+^:CD8^+^ T cell ratios, indicating redundancy. Thus, CD8^+^ T cell frequency was used as the representative variable.

Based on these findings, CD8^+^ T cell frequency, and HLA-DR and CD69 expression on circulating CD4^+^ T cells were selected for further evaluation as candidate prognostic biomarkers (**Figure 2**), as these were the only non-redundant immune variables significantly or near-significantly associated with clinical outcomes in the univariable analysis. HLA-DR expression on CD4^+^ T cells and CD8^+^ T cell frequency showed potential discriminatory ability for 2-year OS (AUC = 0.674, *p* = 0.036 and AUC = 0.687, *p* = 0.024, respectively), while CD69 expression on CD4^+^ T cells had weaker prognostic value (AUC = 0.623, *p* = 0.140) (**Figure 2A, D, G**). When patients were stratified for Kaplan-Meier analysis, those with high HLA-DR expression on CD4^+^ T cells had significantly longer OS compared to those with low expression; however, this association did not retain significance in the multivariable model after adjusting for confounding factors (HR = 0.948, *p* = 0.140) (**Figure 2B−C**). Similarly, patients with high CD69 expression on CD4^+^ T cells showed a trend toward longer OS, approaching significance in the multivariable model (HR = 0.866, *p* = 0.051) (**Figure 2E−F**). Conversely, increasing circulating CD8^+^ T cell frequency remained independently associated with worse OS (HR = 1.064, *p* = 0.002) (**Figure 2H−I**). Regarding EFS, only HLA-DR expression on CD4^+^ T cells was further examined (**Figure 2J−L**), as it was the only immune variable significantly associated with progression/recurrence in the univariable Cox regression. HLA-DR expression on CD4^+^ T cells showed potential to predict progression/recurrence (AUC = 0.732, *p* = 0.004), with high levels of this molecule significantly associated with longer EFS (HR = 0.432, *p* = 0.030), independently from confounders (HR = 0.914, *p* = 0.026).

**Figure 2.**
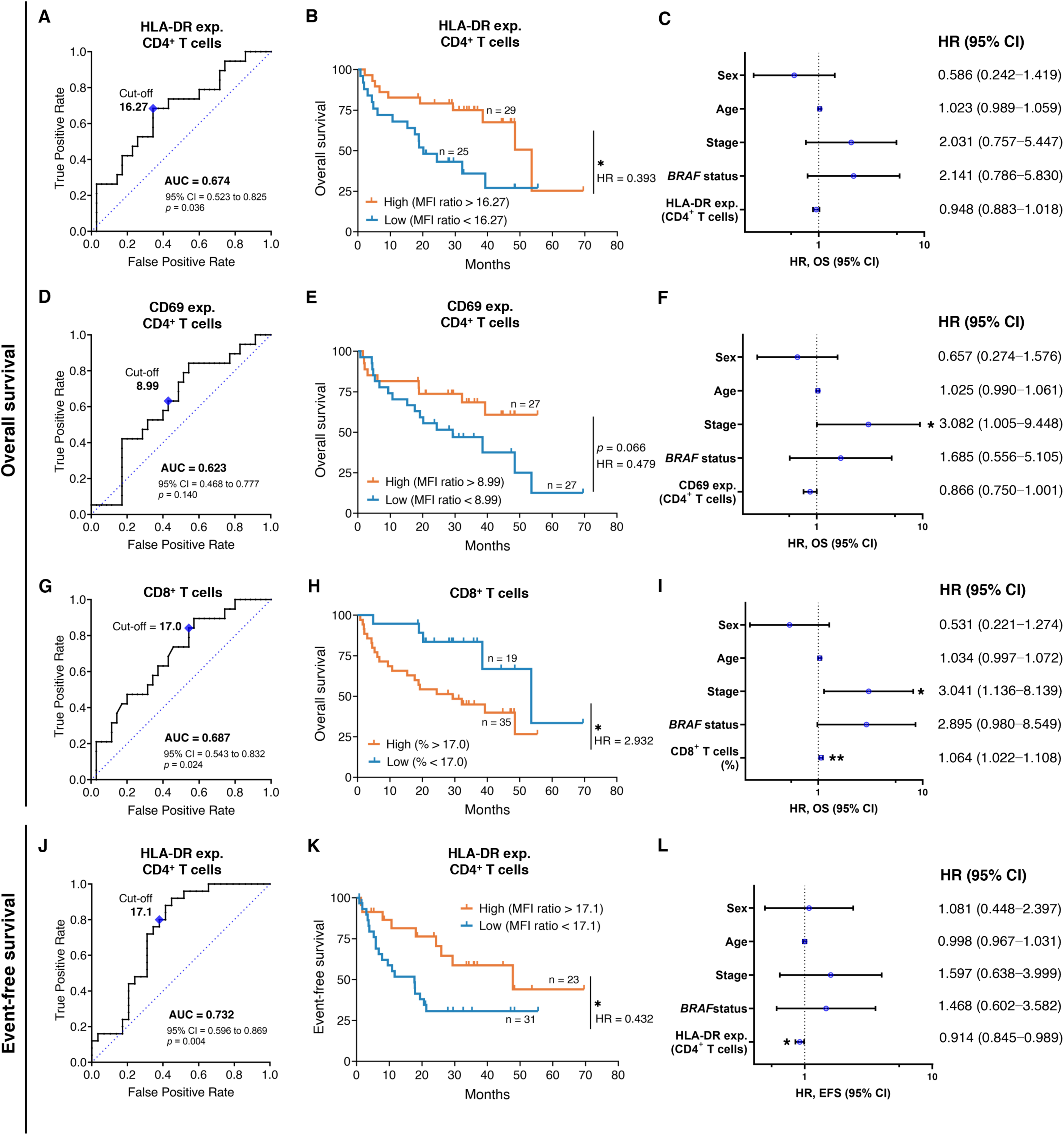
Prognostic value of circulating immune biomarkers in cutaneous melanoma. Flow cytometry-based immune variables from PBMCs of 54 treatment-naïve cutaneous melanoma patients were evaluated for association with OS and EFS. HLA-DR and CD69 expression was defined as MFI ratios between marker-positive and -negative populations. ROC curves were built to assess the discriminatory performance of each biomarker based on outcome status at the 2-year timepoint (left panels). Patients were stratified into high and low groups using optimal cut-off values (maximized Youden’s index) for Kaplan-Meier survival analysis with log-rank testing (middle panels). Multivariable Cox regression was performed on continuous variables, adjusted for sex (reference: male), age, stage (reference: stage III), and *BRAF* mutational status (reference: wildtype). Two patients were excluded from multivariable models due to missing clinical data (right panels). (**A−C**) HLA-DR expression on CD4^+^ T cells, (**D−F**) CD69 expression on CD4^+^ T cells, and (**G−I**) frequency of circulating CD8^+^ T cells (gated on live PBMCs), in relation to OS. (**J−L**) HLA-DR expression on CD4^+^ T cells in relation to EFS. Abbreviations: AUC, area under the curve; CI, confidence intervals; exp., expression; EFS, event-free survival; HR, hazard ratio; MFI, median fluorescence intensity; OS, overall survival; PBMCs, peripheral blood mononuclear cells. *, *p* < 0.05; **, *p* < 0.01.

To assess combined prognostic value (**Figure 3**), we constructed a multivariable model incorporating a clinical risk score (sex, age, stage, *BRAF* status) with HLA-DR and CD69 expression on CD4^+^ T cells, and CD8^+^ T cell frequency (**Figure 3A**, Supplementary Table S7). Increasing clinical risk score and CD8^+^ T cell frequency were associated with poor OS (HR = 3.017, *p* = 0.004 and HR = 1.044, *p* = 0.035), while increasing CD69 expression on CD4^+^ T cells predicted improved OS (HR = 0.819, *p* = 0.047). HLA-DR expression on CD4^+^ T cells showed a trend towards longer OS (HR = 0.952, *p* = 0.259). Overall, the model robustly predicted 2-year OS (AUC = 0.872, *p* < 0.0001), outperforming the clinical risk score alone (**Figure 3B**). Stratification using the optimal ROC cut-off (89.5% sensitivity, 72.7% specificity) identified high- and low-risk groups with significantly different OS (HR = 6.091, *p* < 0.0001; **Figure 3C**). The final EFS prognostic model only included the clinical risk score and HLA-DR expression on CD4^+^ T cells (**Figure 3D**, Supplementary Table S7). Increasing HLA-DR expression was significantly associated with delayed progression (HR = 0.919, *p* = 0.040), whereas the clinical risk score was not (HR = 1.549, *p* = 0.428). Despite moderate performance, the model demonstrated significant discrimination value (AUC = 0.763, *p* = 0.001), outperforming the clinical score alone, and identified high-risk patients with shorter EFS with 75% sensitivity and 71.4% specificity (HR = 2.284, *p* = 0.027; **Figure 3E−F**). Stage-stratified analysis confirmed that both OS and EFS models effectively separated risk groups, particularly in stage IIC/III CM (Supplementary Figure S7).

**Figure 3.**
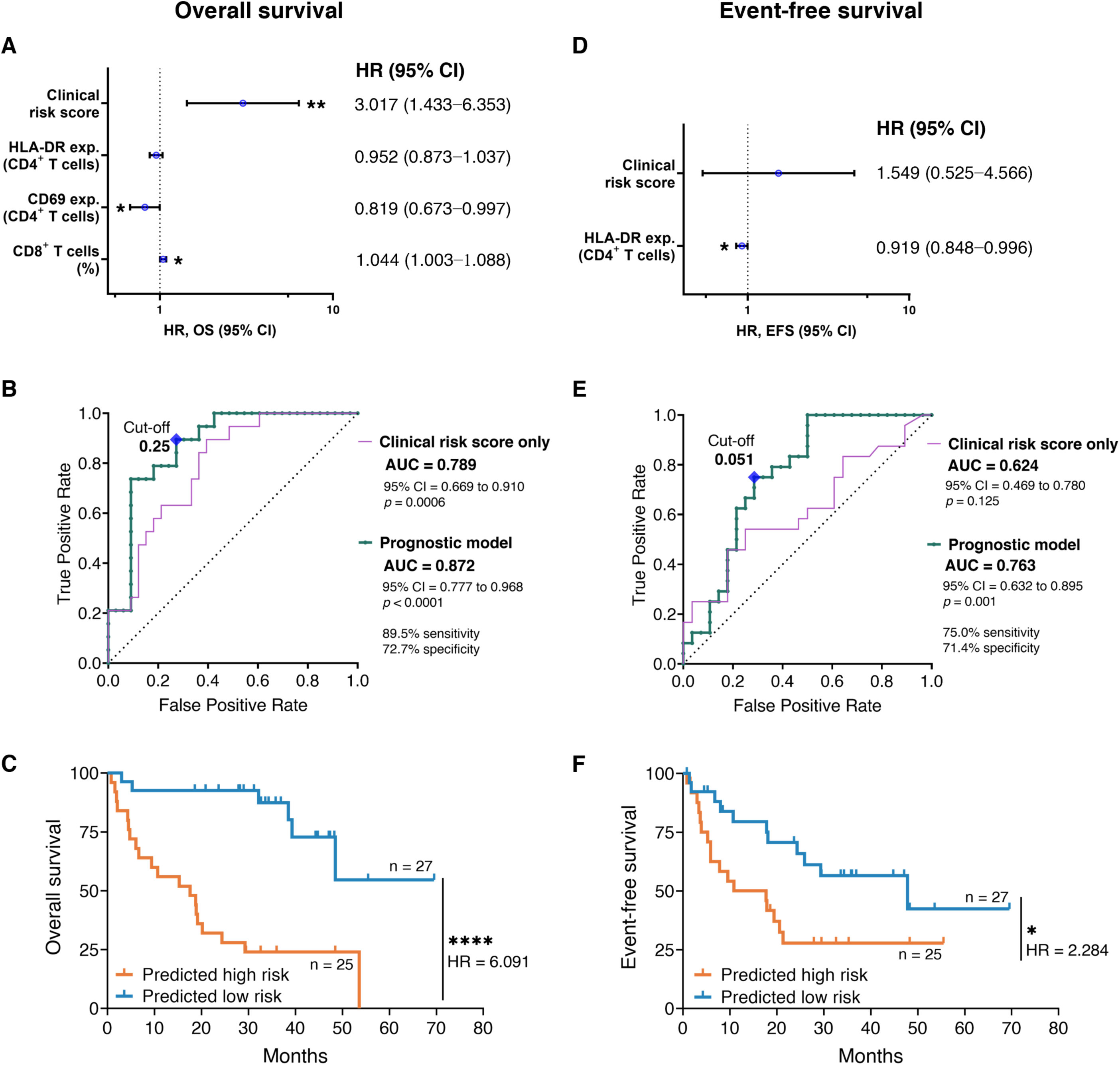
Final prognostic Cox models identify circulating immune biomarkers independently associated with survival and progression/recurrence in cutaneous melanoma. Multivariable Cox regressions were performed combining selected flow cytometry-based immune variables from PBMCs of 52 treatment-naïve cutaneous melanoma patients with calculated clinical risk scores (top panels). HLA-DR and CD69 expression was defined as MFI ratios between marker-positive and -negative populations. The clinical risk score corresponds to the linear predictor from a Cox regression including sex, age, stage, and *BRAF* status. ROC curves were built to assess the discriminatory performance of each model based on its linear predictors (middle panels). Patients were stratified into high- and low-risk groups using optimal cut-off values (maximized Youden’s index) for Kaplan-Meier survival analysis with log-rank testing (bottom panels). Prognostic models are shown for (**A−C**) OS and (**D−F**) EFS.

Together, these results support the prognostic relevance of circulating immune markers. CD69 and HLA-DR expression on CD4^+^ T cells and CD8^+^ T cell frequency may serve as informative and accessible biomarkers for risk stratification in CM.

### 3.3. Elevated HLA-DR and CD69 expression on CD4^+^ T cells as potential baseline predictors of ICI benefit in CM

We next evaluated whether any of the previously assessed baseline circulating immune features could predict therapeutic benefit in 43 ICI-treated patients. Baseline immune subset frequencies did not differ between benefit groups (Supplementary Figure S8). However, baseline HLA-DR and CD69 expression on circulating CD4^+^ T cells tended to be lower in patients without therapeutic benefit (**Figure 4A, B**). No significant differences were observed for the remaining activation or polarization markers (**Figure 4C−M**).

**Figure 4.**
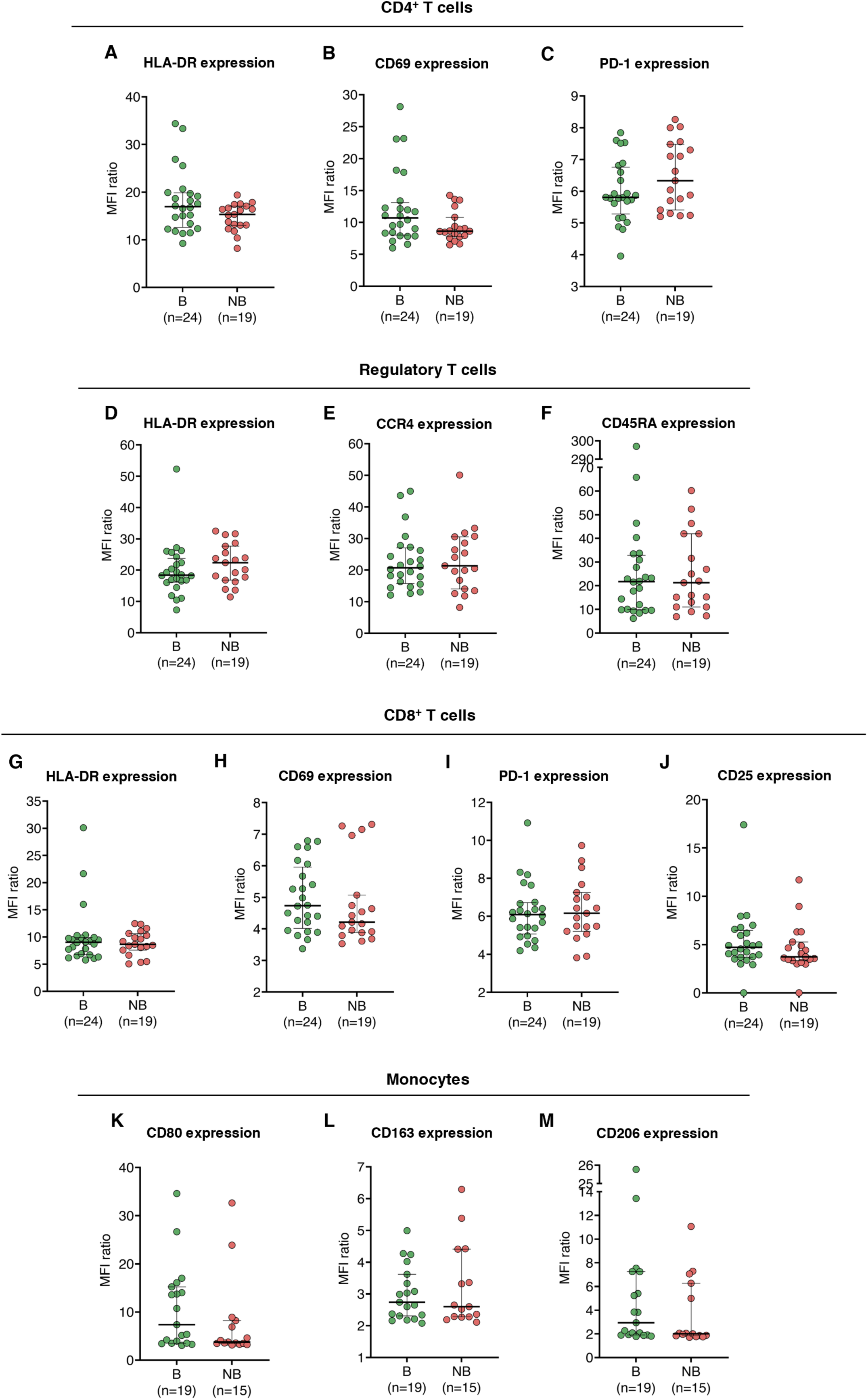
Baseline expression of immune functional markers is comparable between cutaneous melanoma patients stratified by therapeutic benefit from ICIs. Expression of functional markers was quantified by flow cytometry in PBMCs from 43 CM patients prior to receiving ICIs, stratified by therapeutic benefit (yes vs. no). Expression levels are defined as the MFI ratio between marker-positive and -negative populations. Analysed markers include (**A**) HLA-DR, (**B**) CD69, and (**C**) PD-1 on CD4^+^ T cells; (**D**) HLA-DR, (**E**) CCR4, and (**F**) CD45RA on Tregs; (**G**) HLA-DR, (**H**) CD69, (**I**) PD-1, and (**J**) CD25 on CD8^+^ T cells; (**K**) CD80, (**L**) CD163, and (**M**) CD206 on monocytes. Each dot represents one patient. Data are shown as median with interquartile range (IQR). Statistical comparisons were performed using the Mann-Whitney U test. Abbreviations: B, therapeutic benefit; CM, cutaneous melanoma; ICIs, immune-checkpoint inhibitors; MFI, median fluorescence intensity ratio; NB, no therapeutic benefit; PBMCs, peripheral blood mononuclear cells.

To complement these findings, univariable logistic regression analysis was performed to evaluate the predictive value of baseline immune markers and key clinicopathological variables. Therapeutic benefit (yes/no) was used as the binary outcome (Supplementary Table S8). No variable reached statistical significance; however, higher expression of HLA-DR (OR = 1.155, *p* = 0.076) and CD69 (OR = 1.189, *p* = 0.080) on circulating CD4^+^ T cells showed trends toward predicting benefit. ROC curves revealed modest, not statistically significant discriminatory power for both markers (**Figure 5A, B**; AUC = 0.640, *p* = 0.118 and AUC = 0.632, *p* = 0.142). Nevertheless, using optimal cut-off values calculated by maximizing Youden’s index, HLA-DR expression on CD4^+^ T cells showed a trend toward association with therapeutic benefit that did not reach statistical significance (**Figure 5C**), whereas high CD69 expression was significantly associated with benefit, with a greater proportion of patients in the high-expression group achieving benefit compared with the low-expression group (**Figure 5D**). In the multivariable analysis, HLA-DR expression on CD4^+^ T cells emerged as an independent predictor of therapeutic benefit (**Figure 5E**; OR = 1.206, *p* = 0.044), while CD69 expression was not independently predictive (**Figure 5F**; OR = 1.168, *p* = 0.106).

**Figure 5.**
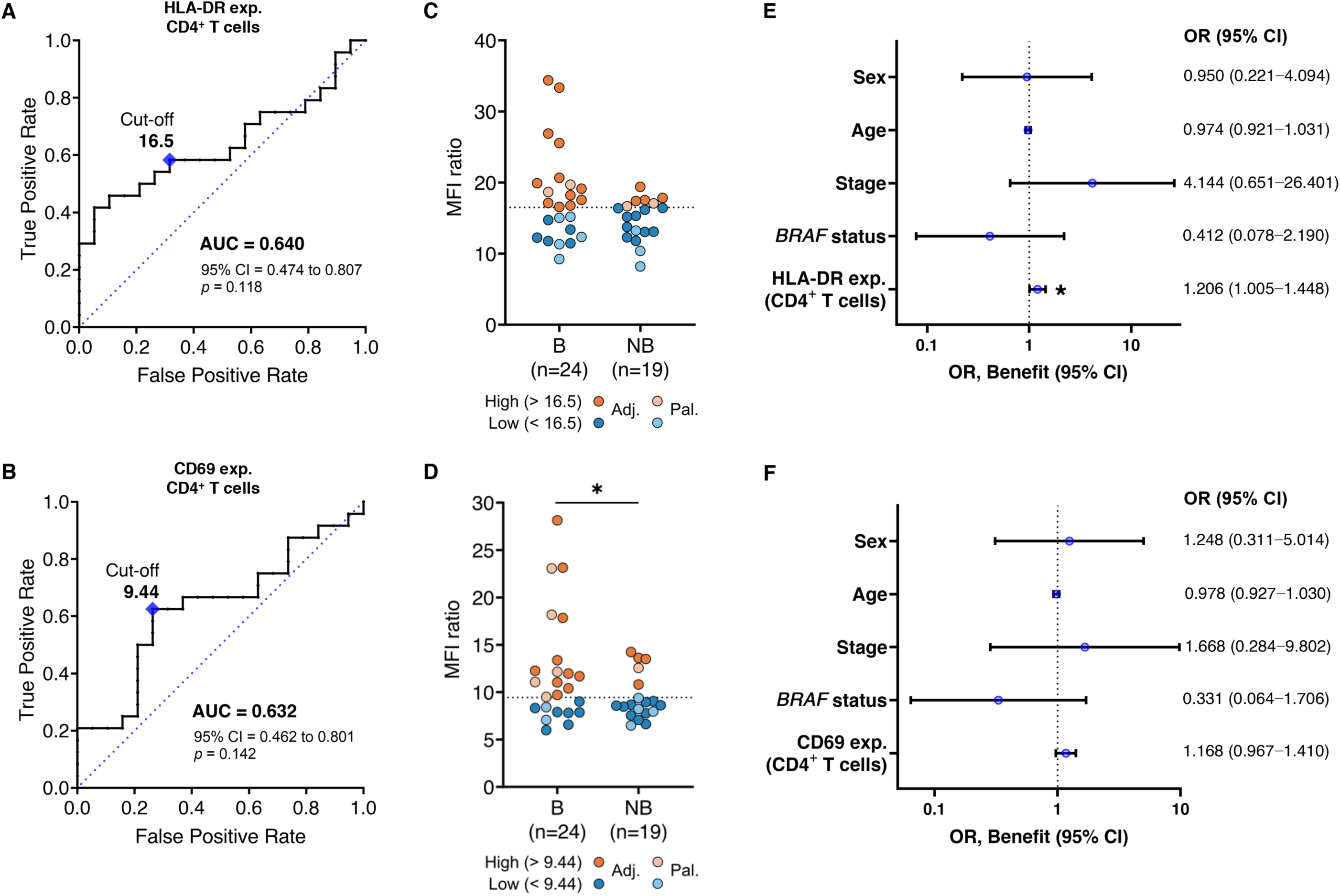
Predictive value of HLA-DR or CD69 expression on CD4^+^ T cells in CM. Flow cytometry-based immune variables from PBMCs of 43 CM patients were assessed at baseline for association with therapeutic benefit following ICI. Expression was defined as MFI ratios between marker-positive and -negative populations. (**A−B**) ROC curves were built to assess the predictive performance of each biomarker using therapeutic benefit as the binary outcome. (**C−D**) Patients were stratified into high (orange) and low (blue) expression groups using optimal cut-off values (maximized Youden’s index) for Fisher’s exact test comparisons; patients treated in the palliative setting are represented in a lighter hue. (**E−F**) Multivariable logistic regression was performed on continuous expression values, adjusted for sex (reference: male), age, stage (reference: stage III), and *BRAF* mutational status (reference: wildtype). Two patients were excluded from multivariable models due to missing clinical data. Each dot represents a patient. Top panels: HLA-DR expression on CD4^+^ T cells. Bottom panels: CD69 expression on CD4^+^ T cells. Abbreviations: Adj., adjuvant; AUC, area under the curve; B, therapeutic benefit; CI, confidence intervals; CM, cutaneous melanoma; exp., expression; ICI, immune-checkpoint inhibition; MFI, median fluorescence intensity; NB, no therapeutic benefit; OR, odds ratio; Pal., palliative; PBMCs, peripheral blood mononuclear cells. *, *p* < 0.05.

A combined logistic model incorporating a composite clinical risk score with both markers (**Figure 6**, Supplementary Table S9) did not yield statistically significant variables (**Figure 6A**). However, the model achieved moderate predictive performance of ICI benefit (AUC = 0.775, *p* = 0.0027), outperforming the clinical risk score alone, which lacked predictive value (**Figure 6B**). Using the optimal ROC cut-off (65.2% sensitivity, 88.9% specificity), a greater proportion of patients in the high-predicted-probability group achieved therapeutic benefit compared with the low-predicted-probability group (**Figure 6C**).

**Figure 6.**
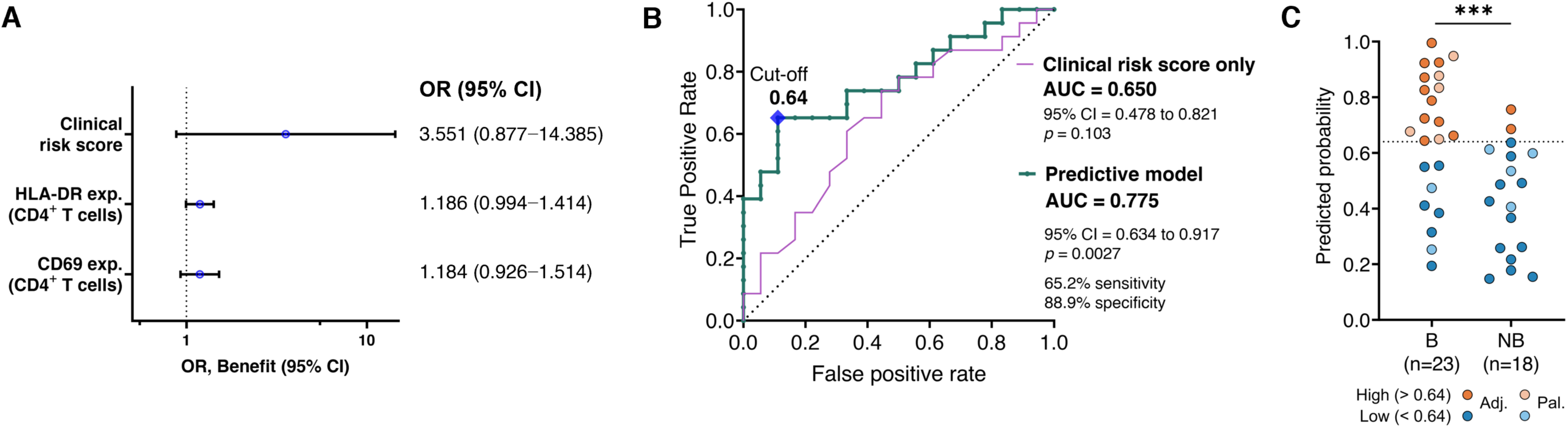
Final predictive logistic regression model highlights baseline circulating immune biomarkers potentially associated with therapeutic benefit from ICIs in CM. (**A**) Multivariable logistic regression was performed using flow cytometry data from PBMCs of 41 cutaneous melanoma patients collected prior to ICI initiation, with therapeutic benefit as the binary outcome. The model included HLA-DR and CD69 expression on CD4^+^ T cells (defined as MFI ratios between marker-positive and -negative populations) along with a clinical risk score. The clinical risk score was calculated as the log-odds from a separate logistic regression incorporating sex, age, stage, and *BRAF* mutational status. (**B**) A ROC curve was generated to evaluate the predictive performance of the model based on its predicted probabilities. (**C**) Patients were stratified into biomarker-high (orange) and -low (blue) groups using the optimal probability cut-off (maximized Youden’s index) for Fisher’s exact test comparisons; patients treated in the palliative setting are represented in a lighter hue. Each dot represents a patient. Abbreviations: AUC, area under the curve; B, therapeutic benefit; CI, confidence intervals; CM, cutaneous melanoma; exp., expression; ICI, immune-checkpoint inhibitors; MFI, median fluorescence intensity; NB, no therapeutic benefit; OR, odds ratio; PBMCs, peripheral blood mononuclear cells. ***, *p* < 0.001.

Applying the same model to longitudinal samples revealed that predictive reliability declined during therapy (Supplementary Figure S9). Predicted probabilities fluctuated considerably and did not consistently anticipate clinical events.

Taken together, our findings suggest that, while CD4^+^ T cell activation markers, namely HLA-DR and CD69, may hold baseline predictive value, their dynamic monitoring during ICI therapy appears less informative in this cohort.

### 3.4. Tumor immunity as a complementary prognostic tool in CM

Having identified HLA-DR and CD69 expression on circulating CD4^+^ T cells, along with the frequency of circulating CD8^+^ T cells, as potential predictors of clinical outcomes in CM, we next evaluated the relationship between these blood immune markers and tumor-infiltrating immune populations (Supplementary Figure S10). We analyzed 11 paired blood-tumor samples by flow cytometry, with tumor analysis restricted to immune cell subset frequencies due to low immune infiltration in several tumor samples. Circulating HLA-DR^+^ CD4^+^ T cells positively correlated with tumor HLA-DR^+^ and CD69^+^ T cell frequencies (Supplementary Figure S10B−E). However, higher HLA-DR expression on circulating CD4^+^ T cells inversely correlated with the frequency of tumor HLA-DR^+^ and CD69^+^ CD4^+^ T cells (Supplementary Figure S10F−G). CD69 expression on circulating CD4^+^ T cells did not correlate with tumor CD69^+^ T cells, but was positively correlated with CD80^+^ and CD163^+^ macrophage infiltration (Supplementary Figure S10H−I). Circulating CD8^+^ T cell frequency did not correlate with any tumor population (Supplementary Figure S10A). These results suggest that the frequency of activated CD4^+^ T cells in blood may reflect the frequency of activated T cells in the tumor, though marker expression levels on circulating cells do not directly correspond to expression levels in the tumor.

To complement immune profiling, we also analyzed tumor immune-related gene expression in 26 CM samples using a 26-gene panel (**Figure 7**). Unsupervised clustering defined two patient groups (**Figure 7A**) with no significant associations with sex, age, stage, or *BRAF* status (Supplementary Figure S11A−D). With respect to systemic therapy, 12 of the 26 patients did not receive subsequent treatment. Among the remaining 14 patients, 8 were treated with ICIs, and 6 received targeted therapy. As a result, no clear associations could be established between gene expression clusters and therapeutic response. However, patient cluster 2 was enriched for patients with longer OS and EFS and contained a higher proportion of lymph node metastasis samples compared with cutaneous metastases or primary CM, although this difference was not significant (Supplementary Figure S11E). Kaplan-Meier analysis confirmed longer OS and EFS in cluster 2 (**Figure 7B−C**). Differential gene expression analysis identified 13 upregulated genes in patient cluster 2 (*STAT1*, *CXCL9*, *CXCL10*, *IDO1*, *IFNG*, *GZMB*, *PRF1*, *FOXP3*, *CCR2*, *CCR4*, *TGFB*, *HLA-DRA*, and *TNFA*), key drivers of cluster separation (**Figure 7D**). While cluster 2 contained more lymph node samples, a direct comparison by site revealed only *IL12A* as differently expressed between lymph node and cutaneous samples (Supplementary Figure S11F), indicating that the clustering and its prognostic implications reflect underlying gene expression patterns rather than sampling bias.

**Figure 7.**
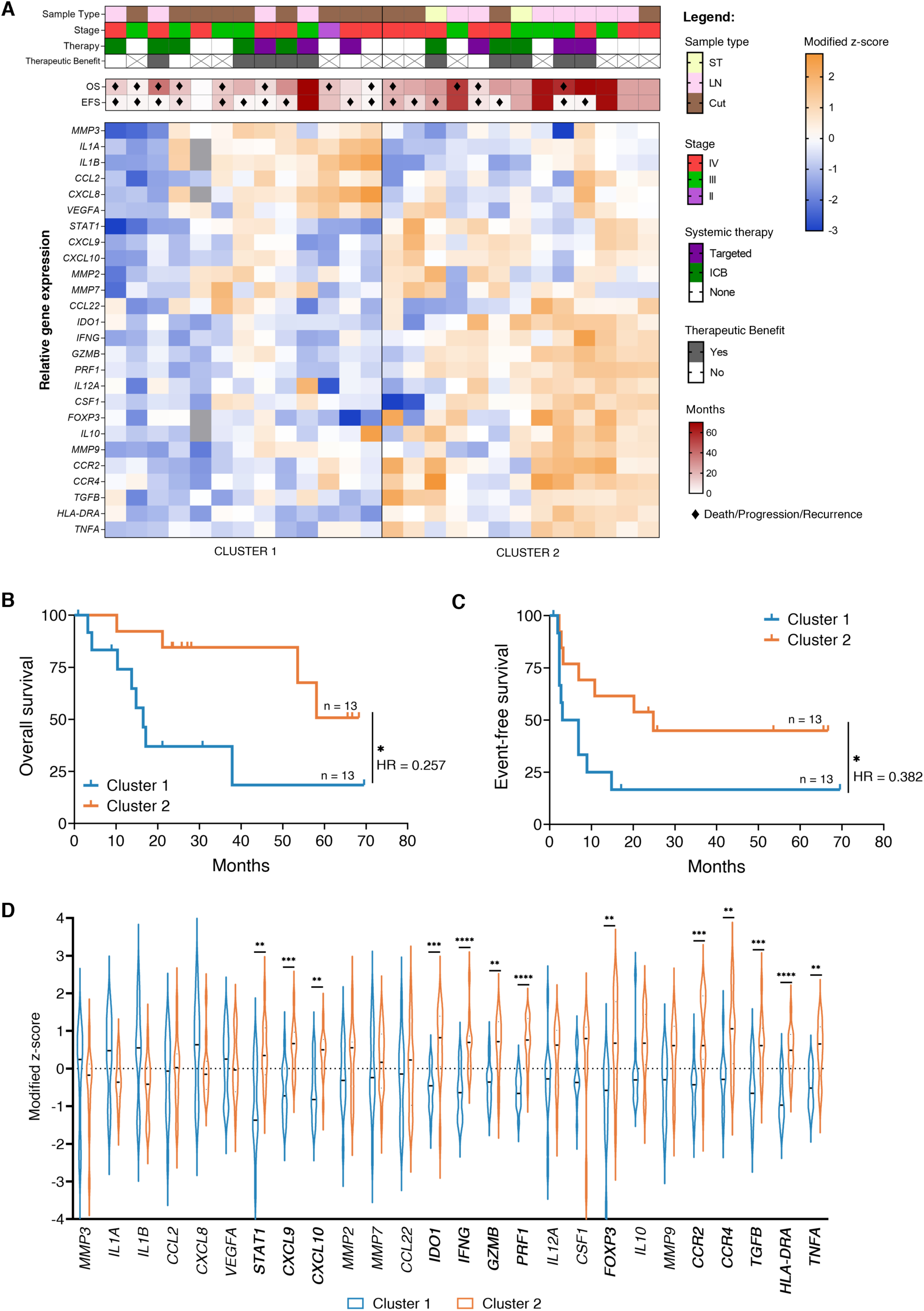
Tumor gene expression profiling stratifies cutaneous melanoma (CM) patients into prognostically distinct clusters. (**A**) Heatmap showing unsupervised hierarchical clustering of 26 tumor samples from CM patients based on a 26-gene immune-related panel. Relative gene expression was quantified by RT-qPCR using the ΔΔCt method [54], and normalized as modified z-scores. Samples were grouped into two major clusters according to overall expression similarity. Each column represents a different patient. Top annotations indicate sample type (ST, soft tissue metastasis; LN, lymph node metastasis; Cut, cutaneous sample, primary or metastasis), disease stage, systemic therapy, therapeutic benefit (if applicable), overall survival (OS) and event-free survival (EFS), in months. Death, progression or recurrence are indicated by the symbol ♦. (**B−C**) Kaplan-Meier survival curves comparing (**B**) OS and (**C**) EFS between the two clusters, with log-rank testing. HR, hazard ratio. **(D)** Violin plots showing the distribution of modified z-scores for each gene across both clusters. Differences between clusters were assessed by multiple Mann-Whitney U tests, followed by false discovery rate correction using the Benjamini-Kieger-Yekutieli two-stage step-up method. *, *p* < 0.05; **, *p* < 0.01; *** *p* < 0.001, ****, *p* < 0.0001.

Together, these findings highlight that tumor immune gene signatures, particularly IFN-γ-related pathways, may provide prognostic information complementary to circulating immune biomarkers. Interestingly, the identified blood biomarkers did not correlate with plasma IFN-γ levels (Supplementary Figure S12), suggesting they influence tumor immunity through alternative cytokines or cellular mechanisms.

## 4. Discussion

The advent of ICIs has transformed the treatment of CM, significantly improving patient survival [6, 8]. Nonetheless, resistance to ICIs remains a major obstacle, and current risk stratification relies almost exclusively on clinicopathological parameters which are imperfect prognostic tools [29]. No circulating biomarker has yet been validated to identify patients at higher risk of progression or to predict ICI benefit.

In this exploratory study, we profiled the circulating immune landscape of CM patients, identifying higher HLA-DR and CD69 expression on CD4^+^ T cells, as well as lower frequency of circulating CD8^+^ T cells, as candidate biomarkers associated with better clinical outcomes in CM. When integrated with a composite clinical risk score, these immune features enhanced prognostic accuracy for both OS and EFS. The inverse prognostic association between circulating CD8^+^ T cell frequency and survival, while counterintuitive given their anti-tumoral role, may reflect redistribution from the periphery to tumor sites, as previously reported in melanoma, where lower circulating but increased intra-tumoral CD8^+^ T cells co-occurred [40]. Although our limited paired blood-tumor analysis (n = 11) did not confirm this correlation, discrepancies between peripheral and intra-tumoral CD8^+^ T cell levels have been documented [50, 51]. Supporting our findings, Neagu et al linked higher circulating CD8^+^ T cell frequencies and a lower CD4^+^:CD8^+^ T cell ratio with advanced CM and worse prognosis [50]. While we did not detect a stage-related increase in CD8^+^ T cells – likely because our comparison was limited to stage III and IV patients only – our findings reinforce the prognostic relevance of circulating CD8^+^ T cells and the CD4^+^:CD8^+^ T cell ratio. None of the CD8^+^ T cell activation markers analyzed were associated with patient outcomes, and no functional correlation with tumor-infiltrating cells was observed, highlighting the need for more comprehensive phenotypic profiling in larger cohorts to determine the prognostic value of specific CD8^+^ T cell subsets.

No prior studies have examined the prognostic relevance of HLA-DR or CD69 expression on circulating CD4^+^ T cells in CM. In hepatocellular carcinoma, higher circulating HLA-DR^+^ T cells correlated with improved PFS [52], while in breast cancer, elevated HLA-DR expression on tumour-infiltrating CD8^+^ T cells was linked to longer PFS, with intra-tumoral and circulating levels directly correlated [53, 54]. Applying the same MFI ratio-based quantification approach, our study extends the prognostic potential of HLA-DR expression to circulating CD4^+^ T cells in CM. Similarly, higher tumoral CD4^+^ CD69^+^ T cells have been linked to better outcomes in head and neck squamous cell carcinoma [55], and CD69 expression characterizes tissue-resident memory T cells associated with good prognosis across multiple cancers [56–58]. Although circulating CD69^+^ T cells differ from these tissue-resident subsets, our results identify CD69 expression on circulating CD4^+^ T cells as a novel prognostic marker in CM. Earlier data reporting worse survival with higher circulating CD4^+^ CD69^+^ T cell frequencies in melanoma [59] likely reflects differences in patient stage, management, and quantification strategies.

Beyond prognosis, baseline HLA-DR and CD69 levels also showed predictive potential for ICI benefit, outperforming a model based solely on clinical parameters. This aligns with growing evidence that systemic CD4^+^ immunity is critical for effective immunotherapy [60]. Circulating effector CD4^+^ subsets, including CD69^+^ cells, have been linked to improved immunotherapy outcomes in melanoma and preclinical models [35], while in non-small cell lung cancer, systemic CD4^+^ immunity is required for PD-1/PD-L1 blockade efficacy [61, 62]. However, previous studies focusing on peripheral CD8^+^ T cells reported limited baseline predictive value due to substantial overlap between responders and non-responders [27, 39, 63–66]. Several have suggested that circulating immune markers may be more informative for treatment monitoring rather than for baseline prediction [39, 66, 67]. Notably, these studies focused only on unresectable CM and did not assess HLA-DR or CD69 expression on CD4^+^ T cells. In our mixed adjuvant/palliative cohort, biomarker variability was greater after treatment initiation, possibly reflecting adaptive immune dynamics. A multi-cancer study also questioned the utility of blood immunophenotyping for monitoring ICI response but was limited by short follow-up [27]. In our analysis, the proposed biomarkers showed predictive value at baseline but were less informative for longitudinal monitoring. Together, these observations highlight the need for standardized sampling approaches and larger cohorts to validate these biomarkers and to better characterize their kinetics during therapy.

Our results are supported by prior work from Krieg et al [40], who reported higher baseline HLA-DR and CD69 expression on circulating T cells in responders to anti-PD-1 therapy, with sustained expression after 12 weeks. Intra-tumoral CD69 has also been linked to favorable ICI response, particularly in EOMES^+^CD69^+^CD45RO^+^ memory T cells [68], as EOMES regulates cytokine and cytotoxic effector programs [69]. Our findings further support the utility of evaluating HLA-DR and CD69 expression on CD4^+^ T cells as baseline predictive biomarkers and extend their relevance to the adjuvant treatment setting. Krieg et al also observed lower circulating CD4^+^ and CD8^+^ T cell frequencies in responders [40]. In our cohort, low CD8^+^ frequency was prognostic for improved OS, but not significantly associated with ICI response.

Beyond circulating markers, tumor gene expression profiling identified 13 upregulated genes associated with improved outcomes, overlapping with established IFN-γ-related ICI response signatures [12, 20–22, 70, 71]. While cohort heterogeneity limited direct associations with ICI response, these genes have been previously linked to better prognosis in stage III melanoma, including in patients receiving targeted therapy [72, 73]. Thus, our findings support the hypothesis that IFN-γ-related gene signatures may serve as prognostic indicators of overall patient outcomes in addition to predictive of ICI benefit. IFN-γ signaling activates STAT1, enhances antigen presentation (HLA-DR), induces checkpoint molecules (PD-L1, IDO1), and upregulates cytotoxic mediators (granzymes, perforin), while recruiting effector and regulatory cells (CXCL9, CXCL10, TNF-α, FOXP3, CCR4) to maintain immune equilibrium [21, 66, 70, 74–76]. The concurrent upregulation of *CCR2* and *TGFB* may reflect monocyte recruitment and immune regulation [77, 78]. Given that activated T cells upregulate HLA-DR, elevated CD4^+^ HLA-DR expression may result from IFN-γ-mediated activation, possibly in synergy with TNF-α [49]. This IFN-γ-dominated landscape thus supports both intrinsic tumor immunogenicity and enhanced ICI responsiveness.

Study limitations include the modest, heterogeneous cohort and incomplete clinicopathological data, particularly regarding Breslow thickness and ulceration, which constrained model refinement. Most patients in the prognostic cohort subsequently received ICIs, complicating distinction between purely prognostic factors and those predictive of ICI response. While circulating CD8^+^ T cell frequency was not linked to ICI response, suggesting a true prognostic role, HLA-DR and CD69 expression may be both prognostic and predictive, or merely predictive of response in this ICI-enriched group. Overall, these exploratory findings require validation in larger, multicentric cohorts with balanced clinical representation.

## 5. Conclusion

We identified circulating CD8^+^ T cell frequency and HLA-DR and CD69 expression on CD4^+^ T cells as promising prognostic and predictive biomarkers in CM. This simple, flow cytometry-based approach offers a clinically feasible alternative to complex and costly immune profiling techniques, such as mass cytometry or single-cell RNA sequencing. We have also validated intra-tumoral IFN-γ-related signaling as being associated with improved prognosis in CM, supporting its relevance as both a mechanistic and biomarker framework. Together, these findings (summarized in **Figure 8**) highlight how systemic and tumor immune features jointly influence patient outcomes. Validation in larger, multicentric cohorts with functional studies is needed to confirm the prognostic and predictive value of these biomarkers and clarify their underlying mechanisms, opening new avenues for improved patient stratification and treatment selection.

**Figure 8.**
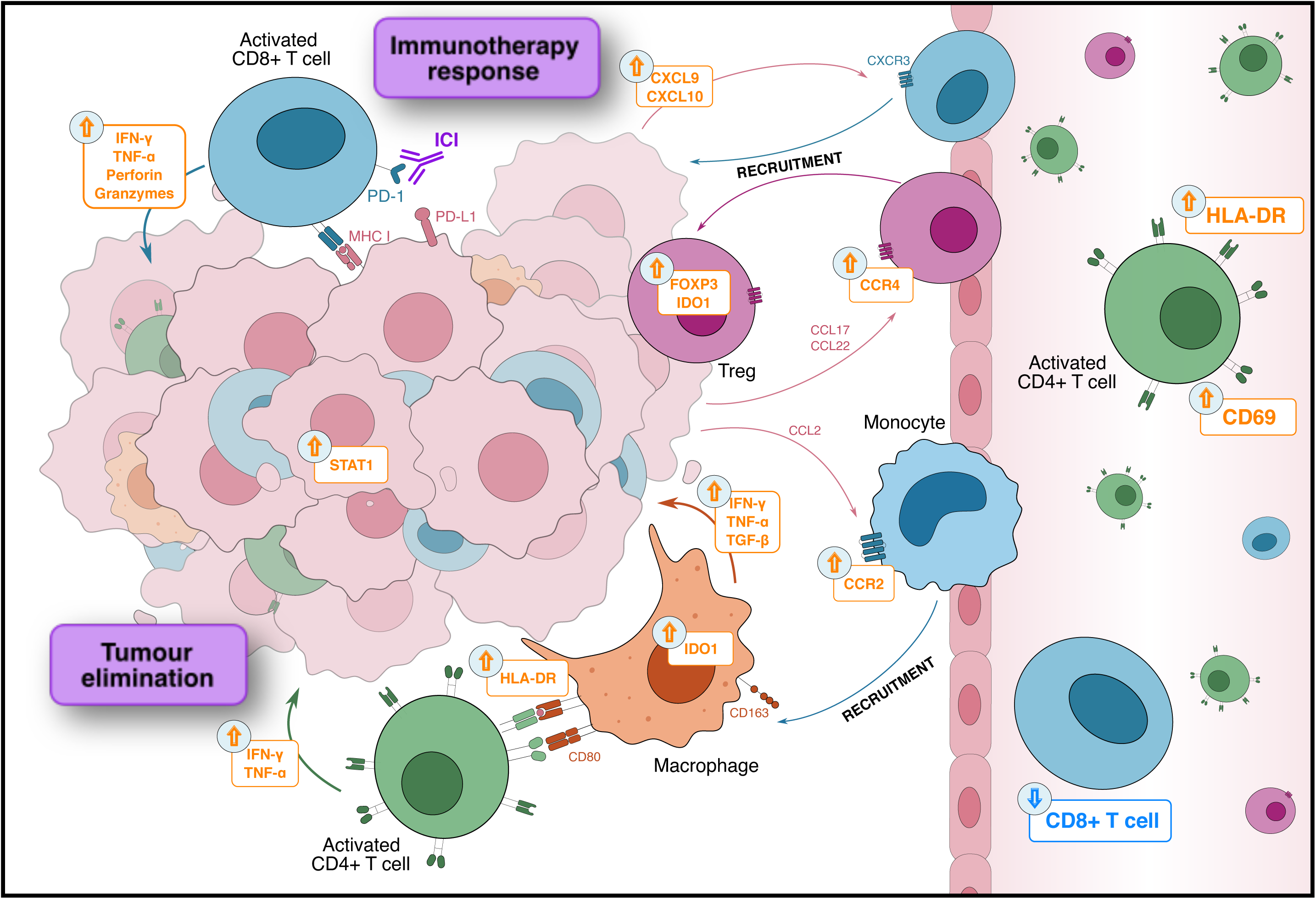
Immune profiling of blood and tumour samples from cutaneous melanoma patients reveals potential prognostic and immunotherapy-response biomarkers. We have identified circulating immune-related biomarkers associated with improved survival and/or response to immune-checkpoint inhibitors (ICIs) that are easily measurable in blood: higher HLA-DR and CD69 expression on CD4^+^ T cells and lower CD8^+^ T cell frequency. In tumor samples from patients with improved survival, we observed activation of IFN-γ-related signaling pathways, previously linked to favorable ICI responses: IFN-γ and TNF-α, produced by activated T cells, activate downstream signaling molecules such as STAT1 in tumor and antigen-presenting cells, leading to upregulation of antigen-presenting molecules (HLA-DR). Activated CD8^+^ T cells also release perforin and granzymes, while chemokines CXCL9 and CXCL10 promote recruitment of additional activated T cells into the tumor microenvironment (TME). Concurrently, negative feedback mechanisms regulating T cell activation are triggered: CCL17 and CCL22 recruit CCR4^+^ regulatory T cells (Tregs, expressing FOXP3), and STAT1 upregulation induces expression of immune checkpoints such as PD-L1 and IDO1. CCL2 produced in the TME recruits monocytes, which can differentiate into tumor-associated macrophages and, in response to immunoregulatory cues, secrete immunosuppressive cytokines such as TGF-β. Together, these mechanisms result in the upregulation of a 13 IFN-γ-related gene signature, associated with improved patient outcomes.

## Funding

This study was supported by Liga Portuguesa Contra o Cancro – Núcleo Regional do Sul – Terry Fox Grant 2020/2021, and by the Research Unit UID/04462: iNOVA4Health—Programme in Translational Medicine and the Associated Laboratory LS4FUTURE (LA/P/0087/2020, https://doi.org/10.54499/LA/P/0087/2020), financially supported by Fundação para a Ciencia e Tecnologia (FCT)/Ministério da Educação, Ciencia e Inovação. AT’s PhD fellowship was funded by FCT (2021.06204.BD, https://doi.org/10.54499/2021.06204.BD).

## Author contributions

A.T.: conceptualization, formal analysis, investigation, writing – original draft; J.M.: investigation, writing – review & editing; H.N. and F.G.: data curation, investigation, resources; R.S.: methodology, writing – review & editing; R.L.: conceptualization, investigation, methodology; C.B.: investigation; D.S.: formal analysis, methodology; E.G., C.P., V.F., E.L.C., C.M., and M.J.P.: investigation, resources; S.C.F.: conceptualization, funding acquisition, project administration, resources, supervision; P.M.P.: data curation, investigation, project administration, resources, supervision, writing – review & editing; M.G.C.: conceptualization, project administration, resources, supervision, writing – review & editing; M.P.: conceptualization, funding acquisition, project administration, supervision, writing – review & editing. All authors approved the final version of the manuscript.

## Data Availability

All data produced in the present study are available upon reasonable request to the authors.

## Acknowledgments

We would like to thank all patients and donors that agreed to participate in this study, all the clinicians at the Surgery, Dermatology, Oncology, Hematology, and Clinical Research Departments at Instituto Português de Lisboa Francisco Gentil, and the Flow Cytometry Facility at NOVA Medical School.

## Declaration of interest statement

The authors declare no conflicts of interest.

## Ethics statement

This study was approved by the Ethics Board Committee from Instituto Português de Oncologia de Lisboa Francisco Gentil E.P.E. (IPOLFG) (UIC/1310) and by the NOVA Medical School Ethics Committee (n° 128/2023/CEFCM), and was conducted in accordance with the Declaration of Helsinki and Portuguese law. Written informed consent was obtained from all participants.

## Data availability statement

The data that support the findings of this study are available from the corresponding author, AT, upon reasonable request.

## Notes

### Competing Interest Statement

The authors have declared no competing interest.

### Author Declarations

Ethics Board Committee from Instituto Português de Oncologia de Lisboa Francisco Gentil E.P.E. (IPOLFG) (UIC/1310) gave ethical approval for this work. Ethics Committee from NOVA Medical School (n. 128/2023/CEFCM) gave ethical approval for this work.

