## Supplementary Information for "Enhanced expression of HLA-DR and CD69 on peripheral CD4^+^ T cells predicts better clinical outcomes in cutaneous melanoma"

Supplementary Table S1. Demographic and clinicopathological characteristics of 54 treatment-naïve CM patients.

| **Variables** | **Stage III CM patients (n = 35)** | **Stage IV CM patients (n = 18)** | **Total (n = 54)** |
| --- | --- | --- | --- |
| Sex, n (%) |  |  |  |
| Male | 18 (51.4) | 14 (77.8) | 33 (61.1) |
| Female | 17 (48.6) | 4 (22.2) | 21 (38.9) |
| Median age, years (range) | 60 (39−84) | 71 (47−91) | 66 (39–91) |
| Stage^†^, n (%) |  |  |  |
| IIC |  |  | 1 (1.9) |
| III |  |  | 35 (64.8) |
| IIIA | 2 (5.7) |  |  |
| IIIB | 11 (31.4) |  |  |
| IIIC | 18 (51.4) |  |  |
| IIID | 4 (11.4) |  |  |
| IV |  |  | 18 (33.3) |
| *BRAF* status, n (%) |  |  |  |
| Wildtype | 19 (54.3) | 17 (94.4) | 36 (66.7) |
| Mutated | 15 (42.8) | 1 (5.6) | 16 (29.6) |
| Missing^‡^ | 1 (2.9) | --- | 2 (3.7) |
| Histology subtype, n (%) |  |  |  |
| Superficial spreading | 16 (45.7) | 9 (50.0) | 26 (48.1) |
| Nodular | 12 (34.3) | 3 (16.67) | 15 (27.8) |
| Acral lentiginous | 1 (2.85) | 3 (16.67) | 4 (7.4) |
| Nevoid | 1 (2.85) | --- | 1 (1.9) |
| Missing^‡^ | 5 (14.3) | 3 (16.67) | 8 (14.8) |
| Median Breslow thickness, mm (range) | 2.8 (0.9−13) | 4.4 (0.9−26) | 3.5 (0.9–26) |
| Missing^‡^ (n, %) | 6 (17.1) | 1 (5.6) | 7 (13.0) |
| Ulceration, n (%) |  |  |  |
| Absent | 15 (42.85) | 9 (50.0) | 25 (46.3) |
| Present | 15 (42.85) | 7 (38.9) | 22 (40.7) |
| Missing^‡^ | 5 (14.3) | 2 (11.1) | 7 (13.0) |
| Median follow-up, months (95% CI) | 35.8 (30.5−41.1) | 44.3 (28.0−60.6) | 36.0 (24.9–47.1) |
| Median OS, months (95% CI) | 48.5 (37.7−59.3) | 15.3 (0.0−39.0) | 39.3 (23.6–55.0) |
| OS status, n (%) |  |  |  |
| Alive | 21 (60.0) | 6 (33.3) | 28 (51.9) |
| Deceased | 14 (40.0) | 12 (66.7) | 26 (48.1) |
| Median EFS, months (95% CI) | 29.4 (8.2−50.6) | 9.6 (1.3−17.9) | 21.3 (11.6–31.0) |
| EFS status, n (%) |  |  |  |
| Progression/disease-free | 17 (48.6) | 7 (38.9) | 25 (46.3) |
| Progression/recurrence | 18 (51.4) | 11 (61.1) | 29 (53.7) |

(continued)

| **Variables** | **Stage III CM patients (n = 35)** | **Stage IV CM patients (n = 18)** | **Total (n = 54)** |
| --- | --- | --- | --- |
| Subsequent therapy, n (%) |  |  |  |
| Targeted therapy | 2 (5.7) | --- | 2 (3.7) |
| Immune-checkpoint inhibitors | 30 (85.7) | 12 (66.7) | 43 (87.0) |
| None | 3 (8.6) | 6 (33.3) | 9 (16.7) |

^†^ Staging was performed according to American Joint Committee on Cancer (AJCC) staging version 8 [49].

^‡^ Data not available in the clinical records or not assessed.

Abbreviations: CM, cutaneous melanoma; EFS, event-free survival; OS, overall survival.

**Supplementary Table S2.** ICI regimens and clinical outcomes in 43 CM patients enrolled while treatment-naïve (baseline).

| **Variables** | **ICI baseline** | | |
| --- | --- | --- | --- |
|  | **Adjuvant (n = 31)** | **Palliative (n = 12)** | **Total (n = 43)** |
| Type of ICI, n (%) |  |  |  |
| Pembrolizumab | 28 (90.3) | 6 (50.0) | 34 (79.1) |
| Nivolumab | 3 (9.7) | 5 (41.7) | 8 (18.6) |
| Nivolumab+ipilimumab | 0 (0) | 1 (8.3) | 1 (2.3) |
| Therapeutic Benefit, n (%) |  |  |  |
| Yes |  |  | 24 (55.8) |
| DFS ≥ 24 months | 17 (54.8) |  |  |
| Good responders |  | 7 (58.3) |  |
| CR |  | 4 (57.1) |  |
| PR |  | 1 (14.3) |  |
| SD > 6 months |  | 2 (28.6) |  |
| No |  |  | 19 (44.2) |
| Recurrence < 24 months | 14 (45.2) |  |  |
| Poor responders |  | 5 (41.7) |  |
| SD < 6 months |  | 0 (0) |  |
| PD |  | 5 (100) |  |

Abbreviations: CM, cutaneous melanoma; CR, complete response; DFS, disease-free survival; ICI, immune‑checkpoint inhibitor; PD, progressive disease; PR, partial response; SD, stable disease.

Supplementary Table S3. Fluorochrome-conjugated anti-human monoclonal antibodies used for flow cytometry-based immunophenotyping of PBMCs and tumor-derived single-cell suspensions**.** All antibodies were obtained from BioLegend.

| **Target antigen** | **Clone** | **Fluorochrome** | **Catalog no.** | **Staining Panel(s)** |
| --- | --- | --- | --- | --- |
| CD3 | HIT3a | PerCP | 300326 | 1, 2 |
| CD4 | OKT4 | FITC | 317408 | 1, 2 |
| CD25 | BC96 | PE | 302606 | 1 |
| CD127 (IL-7Rα) | A019D5 | PE/Cyanine7 | 351320 | 1 |
| HLA-DR | L243 | APC | 307610 | 1, 2 |
| CD194 (CCR4) | L291H4 | Brilliant Violet 421™ | 359414 | 1 |
| CD45RA | HI100 | APC/Cyanine7 | 304128 | 1 |
| CD8a | HIT8a | PE | 300908 | 2 |
| CD69 | FN50 | PE/Cyanine7 | 310912 | 2 |
| CD25 | BC96 | APC/Cyanine7 | 302614 | 2 |
| CD279 (PD-1) | EH12.2H7 | Brilliant Violet 421™ | 329920 | 2 |
| CD45 | HI30 | PerCP | 304026 | 3 |
| CD14 | 63D3 | FITC | 367116 | 3 (PBMCs only) |
| CD16 | 3G8 | Brilliant Violet 421™ | 302038 | 3 (PBMCs only) |
| CD68^†^ | Y1/82A | APC | 333810 | 3 |
| CD80 | 2D10 | FITC | 305206 | 3 (Tumors only) |
| CD80 | 2D10 | PE/Cyanine7 | 305218 | 3 (PBMCs only) |
| CD163 | GHI/61 | PE | 333606 | 3 |
| CD206 (MMR) | 15-2 | APC/Cyanine7 | 321120 | 3 |

^†^Intracellular marker; PBMCS, peripheral blood mononuclear cells.

Supplementary Table S4. Primer sequences used in RT-qPCR**.**

| **Gene** | **Forward primer (5’→3’)** | **Reverse primer (5’→3’)** |
| --- | --- | --- |
| *MMP3* | AGT CTT CCA ATC CTA CTG TTG CT | TCC CCG TCA CCT CCA ATC C |
| *IL1A* | TGG TAG TAG CAA CCA ACG GGA | ACT TTG ATT GAG GGC GTC ATT C |
| *IL1B* | CCT GTC CTG CGT GTT GAA AGA | GGG AAC TGG GCA GAC TCA AA |
| *CCL2* | CAG CCA GAT GCA ATC AAT GCC | TGG AAT CCT GAA CCC ACT TCT |
| *CXCL8* | ACT GAG AGT GAT TGA GAG TGG AC | AAC CCT CTG CAC CCA GTT TTC |
| *VEGFA* | AGG GCA GAA TCA TCA CGA AGT | AGG GTC TCG ATT GGA TGG CA |
| *STAT1* | CGG CTG AAT TTC GGC ACC T | CAG TAA CGA TGA GAG GAC CCT |
| *CXCL9* | CAG CAC CAA CCA AGG GAC TAT C | CAC ATC TGC TGA ATC TGG GTT TAG |
| *CXCL10* | ACC TCC AGT CTC AGC ACC ATG A | TGC AGG TAC AGC GTA CAG TTC T |
| *MMP2* | CTT CAA GGA CCG GTT CAT TTG G | GCC TCG TAT ACC GCA TCA ATC |
| *MMP7* | GGT CAC CTA CAG GAT CGT ATC | CAA CTT TCC TGA AAT GCA GGG G |
| *CCL22* | CCT ACA GAC TGC ACT CCT GGT T | CCT TAT CCC TGA AGG TTA GCA AC |
| *IDO1* | GCT GTT CCT TAC TGC CAA CT | AGC AAA GTG TCC CGT TCT |
| *IFNG* | GTT TTG GGT TCT CTT GGC TGT TA | AAA AGA GTT CCA TTA TCC GCT ACA TC |
| *GZMB* | GGG GGA CCC AGA GAT TAA AA | CCA TTG TTT GGT CCA TAG GAG |
| *PRF1* | GCA ATG TGC ATG TGT CTG TG | GGG AGT GTG TAC CAC ATG GA |
| *IL12A* | CCT TGC ACT TCT GAA GAG ATT GA | ACA GGG CCA TCA TAA AAG AGG T |
| *CSF1* | AGA CCT CGT GCC AAA TTA CAT T | AGG TGT CTC ATA GAA AGT TCG GA |
| *FOXP3* | CAC AAC ATG CGA CCC CCT TTC ACC | AGG TTG TGG CGG ATG GCG TTC TTC |
| *IL10* | GCT GGA GGA CTT TAA GGG TTA CCT | CTT GAT GTC TGG GTC TTG GTT CT |
| *MMP9* | CTG GCA GAG GAA TAC CTG TAC | GAC AGT TGC TTC TGG AGA AGC |

(continued)

| **Gene** | **Forward primer (5’→3’)** | **Reverse primer (5’→3’)** |
| --- | --- | --- |
| *CCR2* | TAC GGT GCT CCC TGT CAT AAA | TAA GAT GAG GAC GAC CAG CAT |
| *CCR4* | AGA AGG CAT CAA GGC ATT TGG | ACA CAT CAG TCA TGG ACC TGA G |
| *TGFB* | CAA GGG CTA CCA TGC CAA CT | AGG GCC AGG ACC TTG CTG |
| *HLA-DRA* | AGT CCC TGT GCT AGG ATT TTT CA | ACA TAA ACT CGC CTG ATT GGT C |
| *TNFA* | CCC CAG GGA CCT CTC TCT AAT C | GGT TTG CTA CAA CAT GGG CTA CA |
| *TBP* | GAG CTG TGA TGT GAA GTT TCC | TCT GGG TTT GAT CAT TCT GTA G |

Supplementary Table S5. Spearman’s rank correlation of CM patient age with circulating immune cell subsets and their functional marker expression **(exp.).** N = 54 (monocyte subset analysis was limited to 41 patients due to low monocyte frequencies).

| **Age vs.** | **Spearman’s r** | ***p*-value** |
| --- | --- | --- |
| CD3^+^ T cells (%) | 0.186 | 0.178 |
| CD4^+^ T cells (%) | 0.106 | 0.446 |
| HLA-DR exp. (CD4^+^ T cells) | −0.280 | 0.040 |
| CD69 exp. (CD4^+^ T cells) | −0.012 | 0.933 |
| PD-1 exp. (CD4^+^ T cells) | −0.316 | 0.020 |
| Regulatory T cells (%) | 0.008 | 0.955 |
| HLA-DR exp. (Tregs) | −0.088 | 0.527 |
| CCR4 exp. (Tregs) | −0.176 | 0.203 |
| CD45RA exp. (Tregs) | −0.259 | 0.059 |
| CD8^+^ T cells (%) | 0.156 | 0.261 |
| Ratio CD4^+^ T:CD8^+^ T | −0.056 | 0.687 |
| Ratio CD8^+^ T:Tregs | 0.007 | 0.961 |
| HLA-DR exp. (CD8^+^ T cells) | −0.261 | 0.057 |
| CD69 exp. (CD8^+^ T cells) | 0.203 | 0.141 |
| PD-1 exp. (CD8^+^ T cells) | −0.079 | 0.572 |
| CD25 exp. (CD8^+^ T cells) | −0.070 | 0.617 |
| Total monocytes (%) | −0.354 | 0.007 |
| Classical monocytes (%) | −0.222 | 0.164 |
| Intermediate monocytes (%) | 0.021 | 0.895 |
| Non-classical monocytes (%) | 0.175 | 0.274 |
| CD80 exp. (monocytes) | 0.190 | 0.233 |
| CD163 exp. (monocytes) | −0.047 | 0.768 |
| CD206 exp. (monocytes) | 0.200 | 0.211 |

Supplementary Table S6. Univariable Cox regression analysis for overall and event-free survival in 54 treatment‑naïve CM patients **(monocyte subsets, n = 41).**

|  | **Overall survival** | | | | **Event-free survival**^¶^ | | | |
| --- | --- | --- | --- | --- | --- | --- | --- | --- |
| **Variables** | ***p*-value** | **HR** | **95% CI** | | ***p*-value** | **HR** | **95% CI** | |
|  |  |  | **Lower** | **Upper** |  |  | **Lower** | **Upper** |
| Sex | 0.123 | 0.515 | 0.222 | 1.197 | 0.891 | 0.949 | 0.451 | 1.997 |
| Age | **0.025** | **1.037** | **1.004** | **1.070** | 0.206 | 1.018 | 0.990 | 1.047 |
| Stage^†^ | **0.016** | **2.595** | **1.193** | **5.644** | **0.079** | **1.969** | **0.925** | **4.190** |
| *BRAF* mutational status^‡^ | 0.957 | 0.978 | 0.431 | 2.217 | 0.702 | 0.856 | 0.387 | 1.894 |
| Breslow thickness^§^ | 0.338 | 0.957 | 0.876 | 1.046 | 0.143 | 0.934 | 0.852 | 1.023 |
| Ulceration^§^ | 0.579 | 0.782 | 0.329 | 1.860 | 0.456 | 0.743 | 0.341 | 1.621 |
| CD3^+^ T cells (%) | 0.101 | 1.022 | 0.996 | 1.050 | 0.359 | 1.011 | 0.987 | 1.036 |
| CD4^+^ T cells (%) | 0.934 | 0.998 | 0.962 | 1.036 | 0.107 | 1.029 | 0.994 | 1.065 |
| HLA-DR exp. (CD4^+^ T cells) | **0.071** | **0.942** | **0.883** | **1.005** | **0.015** | **0.913** | **0.849** | **0.982** |
| CD69 exp. (CD4^+^ T cells) | **0.090** | **0.899** | **0.795** | **1.017** | 0.228 | 0.945 | 0.862 | 1.036 |
| PD-1 exp. (CD4^+^ T cells) | 0.402 | 0.844 | 0.567 | 1.255 | 0.354 | 1.174 | 0.836 | 1.649 |
| Regulatory T cells (%) | 0.340 | 0.898 | 0.720 | 1.120 | 0.270 | 0.884 | 0.710 | 1.100 |
| HLA-DR exp. (Tregs) | 0.650 | 0.988 | 0.940 | 1.040 | 0.884 | 0.997 | 0.953 | 1.043 |
| CCR4 exp. (Tregs) | 0.595 | 0.986 | 0.935 | 1.039 | 0.498 | 1.014 | 0.974 | 1.056 |
| CD45RA exp. (Tregs) | 0.122 | 0.973 | 0.940 | 1.007 | 0.813 | 1.001 | 0.994 | 1.008 |
| CD8^+^ T cells (%) | **0.010** | **1.049** | **1.011** | **1.087** | 0.815 | 1.004 | 0.969 | 1.041 |
| CD4^+^ T:CD8^+^ T (ratio) | **0.053** | **0.597** | **0.354** | **1.007** | 0.346 | 1.182 | 0.835 | 1.672 |
| CD8^+^ T:Tregs (ratio) | **0.005** | **1.079** | **1.024** | **1.137** | 0.643 | 1.012 | 0.961 | 1.067 |
| HLA-DR exp. (CD8^+^ T cells) | 0.164 | 0.912 | 0.801 | 1.038 | 0.371 | 0.960 | 0.878 | 1.050 |
| CD69 exp. (CD8^+^ T cells) | 0.853 | 0.974 | 0.740 | 1.283 | 0.351 | 1.120 | 0.883 | 1.420 |
| PD-1 exp. (CD8^+^ T cells) | 0.519 | 0.914 | 0.695 | 1.201 | 0.736 | 1.042 | 0.822 | 1.319 |
| CD25 exp. (CD8^+^ T cells) | 0.960 | 1.004 | 0.874 | 1.152 | 0.145 | 0.881 | 0.744 | 1.045 |
| Classical monocytes (%) | 0.968 | 0.999 | 0.932 | 1.070 | 0.905 | 0.997 | 0.941 | 1.055 |
| Intermediate monocytes (%) | 0.703 | 0.967 | 0.814 | 1.149 | 0.580 | 0.963 | 0.843 | 1.101 |
| Non-classical monocytes (%) | 0.787 | 1.013 | 0.923 | 1.112 | 0.604 | 1.022 | 0.941 | 1.109 |
| CD80 exp. (monocytes) | 0.497 | 0.979 | 0.920 | 1.041 | 0.376 | 0.976 | 0.925 | 1.030 |
| CD163 exp. (monocytes) | 0.734 | 0.918 | 0.561 | 1.502 | 0.549 | 1.139 | 0.744 | 1.744 |
| CD206 exp. (monocytes) | 0.784 | 0.985 | 0.884 | 1.098 | 0.984 | 0.999 | 0.919 | 1.086 |

^†^One patient with CM stage IIC excluded; ^‡^Two patients with missing *BRAF* status excluded; ^§^Seven patients with missing Breslow thickness and ulceration excluded. Significant and nearly significant *p*-values are shown in bold (*p* < 0.1); ^¶^Events include recurrence (stage IIC/III) and progression (stage IV).

**Categorical variables:** sex (reference: male); stage (reference: stage III); *BRAF* status (reference: wildtype); ulceration (reference: absent).

Abbreviations: CI, confidence interval; CM, cutaneous melanoma; exp., expression; HR, hazard ratio.

Supplementary Table S7. Coefficients from multivariable Cox regressions used to derive the clinical risk scores and final prognostic models for overall survival (OS) and event-free survival (EFS)**.** Data from 52 treatment‑naïve cutaneous melanoma patients was used (2 excluded patients of the total 54 due to missing clinical data). Clinical risk scores correspond to the linear predictor ($X\beta$), calculated as the weighted sum of mean-centred covariates^†^.

|  | **Covariates** | **Value** | **β, OS** | **Mean, OS** | **β, EFS** | **Mean, EFS** |
| --- | --- | --- | --- | --- | --- | --- |
| Clinical risk score regression | Sex | Male = 0 | −0.477 | 0.385 | 0.050 | 0.392 |
|  |  | Female = 1 |  |  |  |  |
|  | Age | Age (in years) | 0.029 | 64.885 | 0.009 | 65.235 |
|  | Stage | Stage III = 0 | 0.847 | 0.346 | 0.694 | 0.333 |
|  |  | Stage IV = 1 |  |  |  |  |
|  | *BRAF* mutational status | Wildtype = 0 | 0.533 | 0.308 | 0.116 | 0.314 |
|  |  | Mutated = 1 |  |  |  |  |
| Final prognostic models | Clinical risk score | $X\beta$ from previous Cox regression | 1.104 | 0.000 | 0.437 | 0.000 |
|  | HLA-DR exp. (CD4^+^ T cells) | MFI ratio | −0.050 | 17.988 | −0.084 | 18.125 |
|  | CD69 exp. (CD4^+^ T cells) | MFI ratio | −0.199 | 10.290 | --- | --- |
|  | CD8^+^ T cells (%) | % of live PBMCs | 0.043 | 22.812 | --- | --- |

^†^$X\beta=\sum\beta(value-mean)$

Supplementary Table S8. Univariable logistic regression analysis for therapeutic benefit in 43 CM patients who underwent treatment with immune-checkpoint inhibitor **(monocyte subsets, n = 34).**

| **Variables at baseline** | ***p*-value** | **OR** | **95% CI** | |
| --- | --- | --- | --- | --- |
|  |  |  | **Lower** | **Upper** |
| Sex | 0.759 | 0.825 | 0.241 | 2.822 |
| Age | 0.458 | 0.983 | 0.941 | 1.028 |
| Stage^†^ | 0.331 | 2.000 | 0.494 | 8.089 |
| *BRAF* mutational status^‡^ | 0.236 | 0.437 | 0.111 | 1.720 |
| Breslow thickness^§^ | 0.139 | 1.120 | 0.964 | 1.302 |
| Ulceration^§^ | 0.634 | 1.358 | 0.385 | 4.787 |
| CD3^+^ T cells (%) | 0.980 | 0.999 | 0.956 | 1.044 |
| CD4^+^ T cells (%) | 0.669 | 0.986 | 0.924 | 1.052 |
| **HLA-DR exp. (CD4^+^ T cells)** | **0.076** | **1.155** | **0.985** | **1.355** |
| **CD69 exp. (CD4^+^ T cells)** | **0.080** | **1.189** | **0.979** | **1.444** |
| PD-1 exp. (CD4^+^ T cells) | 0.119 | 0.611 | 0.328 | 1.136 |
| Regulatory T cells (%) | 0.479 | 1.131 | 0.804 | 1.592 |
| HLA-DR exp. (Tregs) | 0.393 | .965 | 0.888 | 1.048 |
| CCR4 exp. (Tregs) | 0.870 | 0.995 | 0.932 | 1.062 |
| CD45RA exp. (Tregs) | 0.536 | 1.006 | 0.987 | 1.024 |
| CD8^+^ T cells (%) | 0.895 | 1.004 | 0.945 | 1.066 |
| Ratio CD4^+^ T:CD8^+^ T | 0.979 | 1.009 | 0.541 | 1.880 |

(continued)

| **Variables at baseline** | ***p*-value** | **OR** | **95% CI** | |
| --- | --- | --- | --- | --- |
|  |  |  | **Lower** | **Upper** |
| Ratio CD8^+^ T:Tregs | 0.660 | 0.981 | 0.900 | 1.069 |
| HLA-DR exp. (CD8^+^ T cells) | 0.404 | 1.075 | 0.908 | 1.272 |
| CD69 exp. (CD8^+^ T cells) | 0.312 | 1.299 | 0.782 | 2.159 |
| PD-1 exp. (CD8^+^ T cells) | 0.805 | 0.952 | 0.642 | 1.411 |
| CD25 exp. (CD8^+^ T cells) | 0.422 | 1.103 | 0.869 | 1.400 |
| Total monocytes (%) | 0.631 | 1.021 | 0.938 | 1.111 |
| Classical monocytes (%) | 0.455 | 1.036 | 0.943 | 1.139 |
| Intermediate monocytes (%) | 0.707 | 1.045 | 0.832 | 1.311 |
| Non-classical monocytes (%) | 0.273 | 0.929 | 0.814 | 1.060 |
| CD80 exp. (monocytes) | 0.327 | 1.045 | 0.957 | 1.141 |
| CD163 exp. (monocytes) | 0.548 | 0.820 | 0.428 | 1.569 |
| CD206 exp. (monocytes) | 0.363 | 1.091 | 0.905 | 1.314 |

^†^One patient with CM stage IIC excluded; ^‡^Two patients with missing *BRAF* status excluded; ^§^Four patients with missing Breslow thickness and ulceration excluded. Significant and nearly significant *p*-values (*p* < 0.1) are shown in bold.

**Categorical variables:** sex (reference: male); stage (reference: stage III); BRAF status (reference: wildtype); ulceration (reference: absent).

Abbreviations: CI, confidence interval; CM, cutaneous melanoma; exp., expression; OR, odds ratio.

Supplementary Table S9. Coefficients from multivariable logistic regressions used to derive the clinical risk scores and the final model predicting therapeutic benefit. Data from 41 cutaneous melanoma patients prior to receiving immune-checkpoint inhibitors was used (2 excluded patients of the total 43 due to missing clinical data). Risk scores correspond to the linear predictor ($X\beta$), calculated as the weighted sum of covariates from the logistic model^†^. The predicted probabilities (P) of therapeutic benefit were calculated using the logistic regression equation^‡^, using the corresponding $X\beta$.

|  | **Covariates** | **Value** | **β** | **Intercept (β_0_)** |
| --- | --- | --- | --- | --- |
| Clinical risk score regression | Sex | Male = 0 | 0.194 | 1.862 |
|  |  | Female = 1 |  |  |
|  | Age | Age (in years) | −0.026 |  |
|  | Stage | Stage III = 0 | 0.781 |  |
|  |  | Stage IV = 1 |  |  |
|  | *BRAF* mutational status | Wildtype = 0 | −0.719 |  |
|  |  | Mutated = 1 |  |  |
| Final predictve model | Clinical risk score | $X\beta$ from previous logistic regression | 1.267 | −4.644 |
|  | HLA-DR exp. (CD4^+^ T cells) | MFI ratio | 0.170 |  |
|  | CD69 exp. (CD4^+^ T cells) | MFI ratio | 0.169 |  |

^†^$X\beta=(\sum(\beta\times value))+\beta_{0}$; ^‡^$P= \frac{1}{1+e^{-X\beta}}$

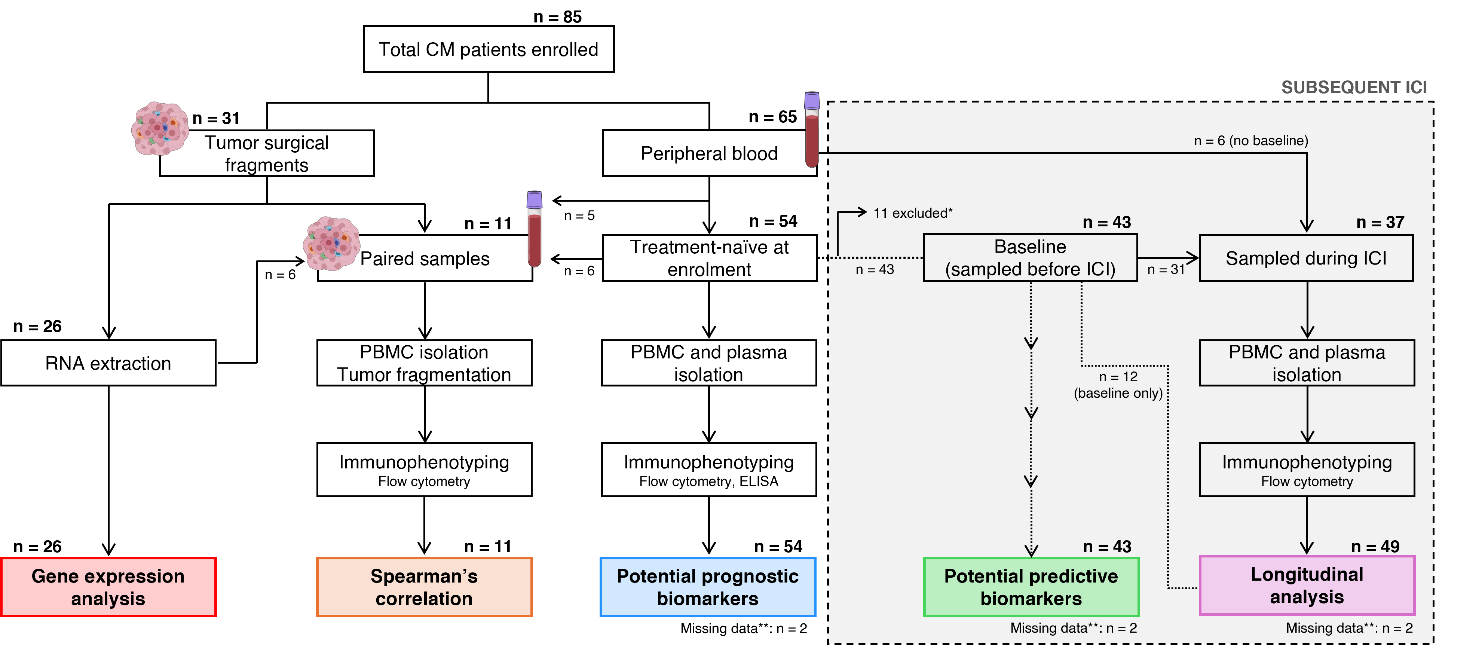

Supplementary Figure S1. Overview of study design and patient distribution. A total of 85 cutaneous melanoma (CM) patients were prospectively enrolled. Tumor surgical fragments from 31 patients enabled gene expression analysis (n = 26) and immune profiling of paired tumor-blood samples (n = 11). Peripheral blood was collected from 65 patients, including 54 treatment‑naïve. Samples were immunophenotyped to identify potential prognostic biomarkers. Of these, 43 patients subsequently initiated immune-checkpoint inhibitors (ICIs), and their baseline samples were used to identify potential predictive biomarkers. Longitudinal blood samples during ICI treatment were available from 37 patients, including 6 without baseline samples; inclusion of 12 baseline-only cases yielded 49 patients for longitudinal immunophenotyping.

Sample sets overlap across analyses, so patient numbers are not mutually exclusive.

*11 patients were excluded due to initiation of targeted therapy or absence of systemic therapy.

**2 patients were excluded from multivariable analyses due to missing *BRAF* status.

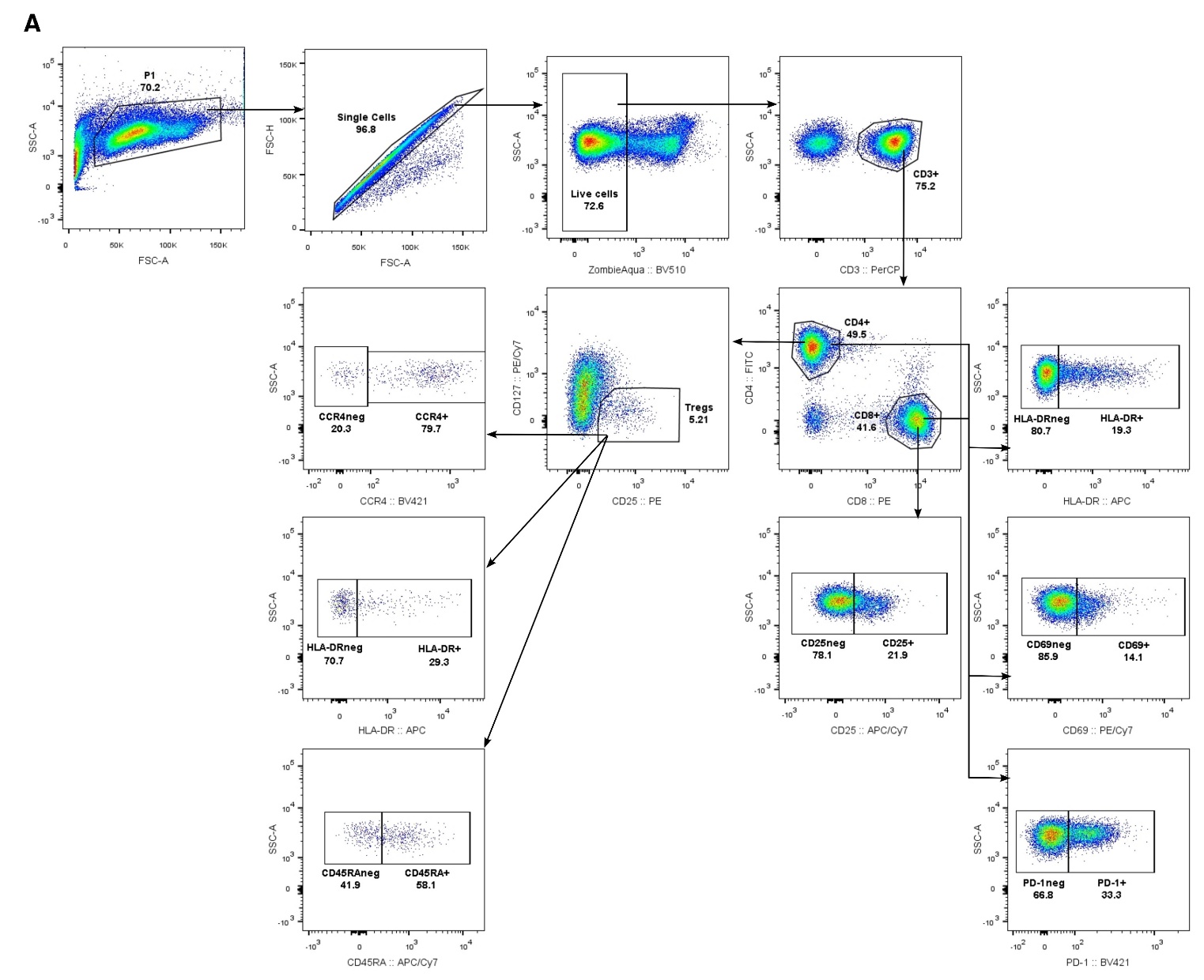

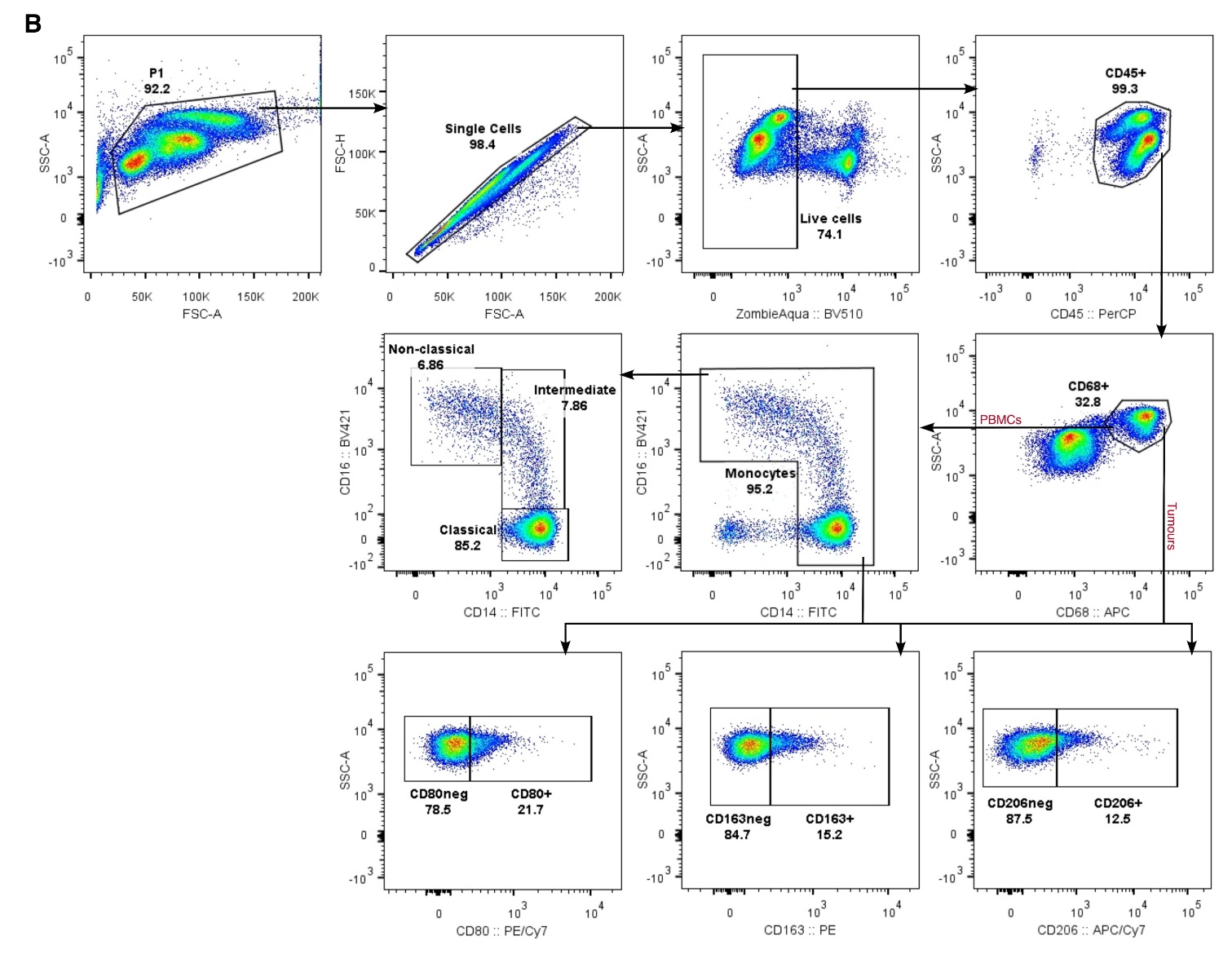
Supplementary Figure S2. Gating strategy for identification of immune cell subsets**.** (**A**) Lymphocyte subsets, including CD4^+^ and CD8^+^ T cell activation markers and markers of Treg immunosuppression. (**B**) Monocyte/macrophage subsets, including M1- and M2-like markers.

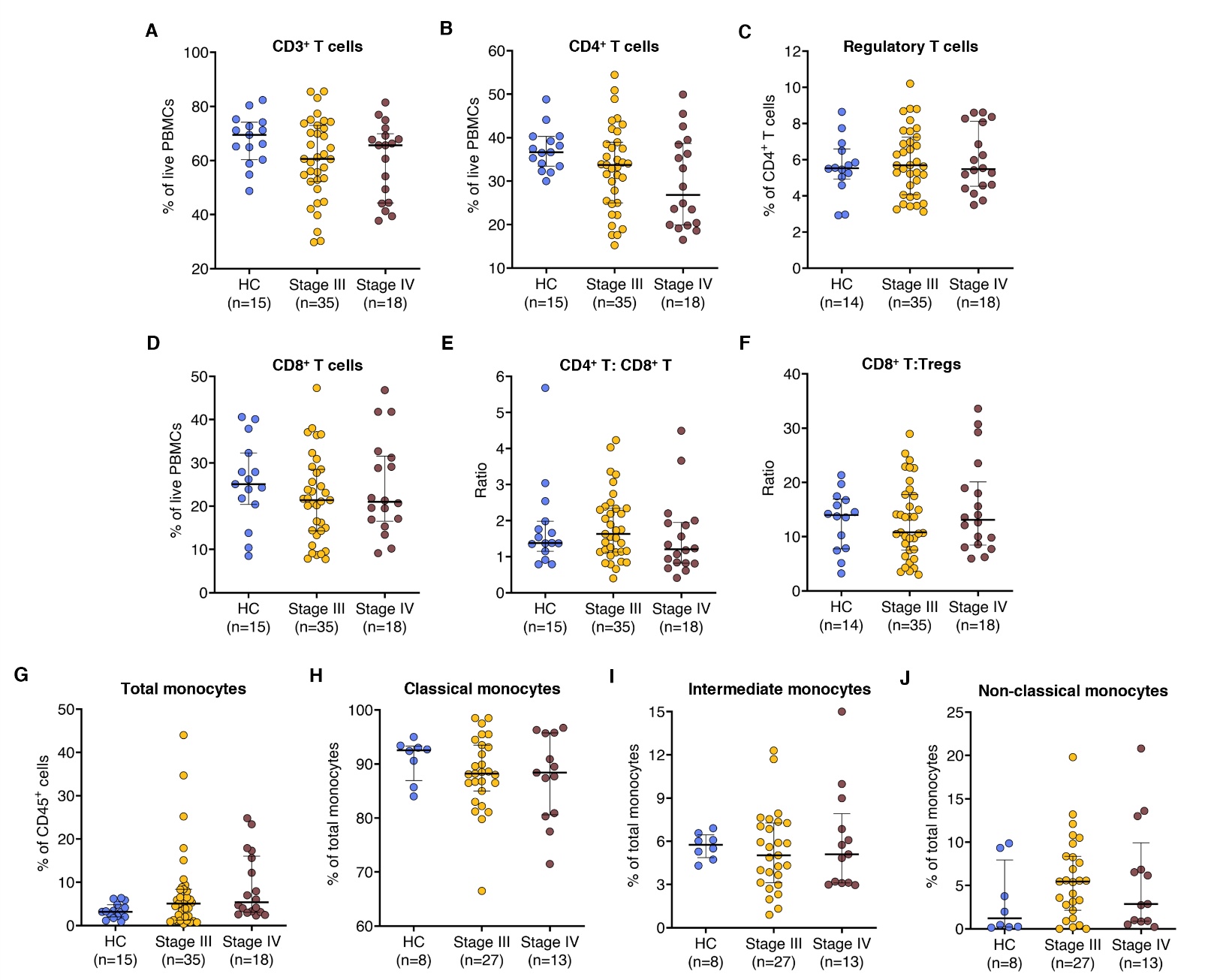
Supplementary Figure S3. Frequencies of circulating immune cell subsets are comparable between healthy donors and cutaneous melanoma patients, and across disease stages**.** Percentages of immune cell subsets were quantified by flow cytometry in peripheral blood mononuclear cells (PBMCs) from 15 healthy controls (HC) and 53 cutaneous melanoma (CM) patients (divided into stage III and stage IV). Analyzed subsets include (**A**) CD3^+^ T cells and (**B**) CD4^+^ T cells (gated on live PBMCs); (**C**) Regulatory T cells (Tregs, gated on CD4^+^ T cells); (**D**) CD8^+^ T cells (gated on live PBMCs); (**E**) CD4^+^:CD8^+^ T cell ratio and (**F**) CD8^+^ T cell:Treg ratio (gated on CD3^+^ T cells); (**G**) total monocytes (gated on CD45^+^ cells); and (**H**) classical, (**I**) intermediate, and (**J**) non-classical monocyte subsets (gated on total monocytes). Monocyte subset analysis was limited to 41 patients and 8 healthy controls due to low monocyte frequencies. Each dot represents an individual. Data are shown as median with interquartile range. Statistical comparisons were performed using the Kruskal‑Wallis test followed by Dunn’s multiple comparisons test.

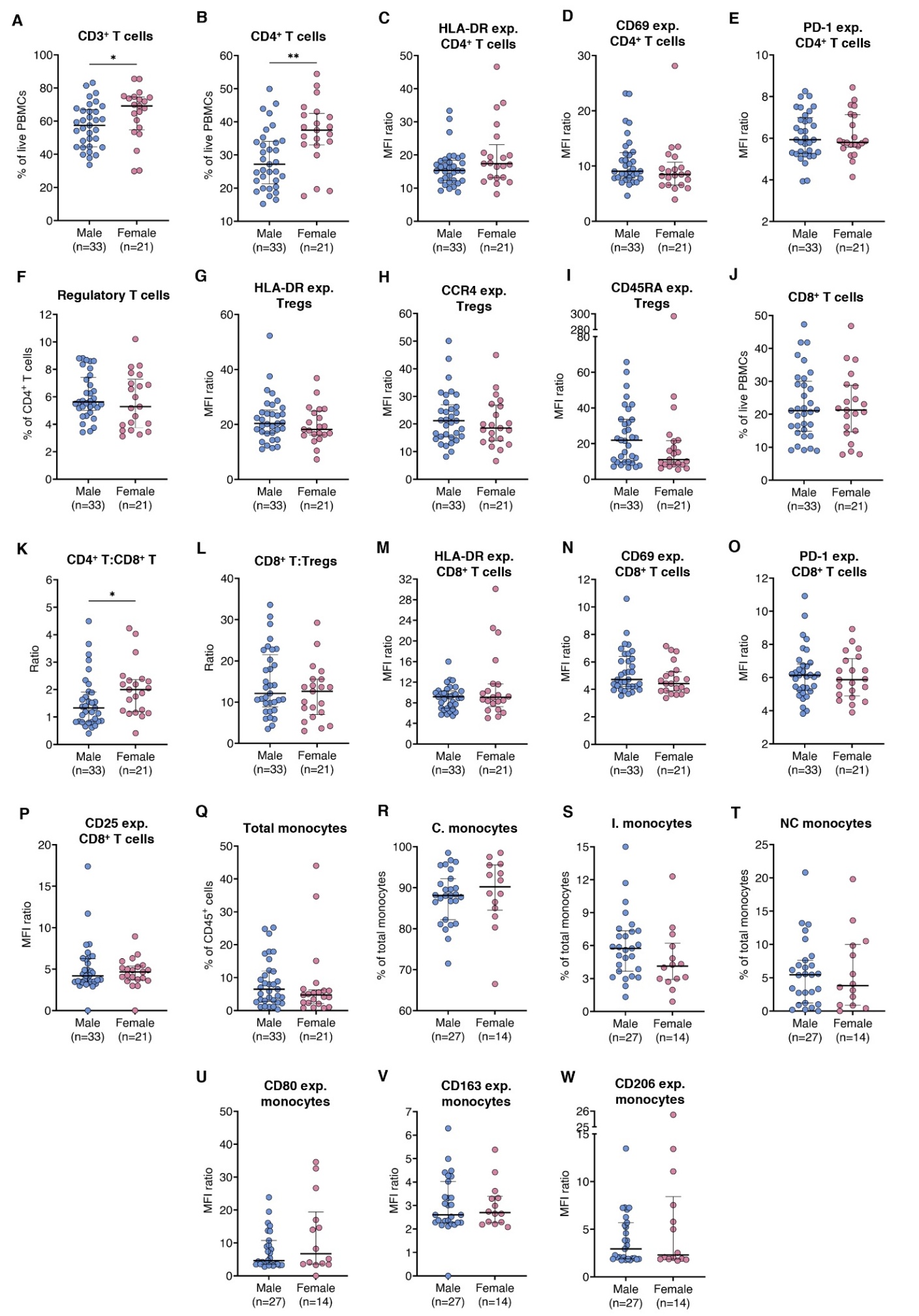
Supplementary Figure S4. Sex-based comparison of circulating immune cell subsets and functional marker expression in cutaneous melanoma patients**.** Flow cytometry was used to quantify immune cell subsets and functional marker expression in peripheral blood mononuclear cells (PBMCs) from 54 cutaneous melanoma (CM) patients, stratified by sex. Marker expression levels (exp.) are defined as the ratio of median fluorescence intensity (MFI) between marker-positive and -negative populations. Analyzed subsets and markers include (**A**) CD3^+^ T cells and (**B**) CD4^+^ T cells (gated on live PBMCs); (**C**) HLA-DR (**D**) CD69, and (**E**) PD1 expression on CD4^+^ T cells; (**F**) Regulatory T cells (Tregs, gated on CD4^+^ T cells); (**G**) HLA‑DR, (**H**) CCR4, and (**I**) CD45RA expression on Tregs; (**J**) CD8^+^ T cells (gated on live PBMCs); (**K**) CD4^+^:CD8^+^ T cell ratio and (**L**) CD8^+^ T cell:Treg ratio (gated on CD3^+^ T cells); (**M**) HLA-DR, (**N**) CD69, (**O**) PD-1, and (**P**) CD25 expression on CD8^+^ T cells; (**Q**) total monocytes (gated on CD45^+^ cells); (**R**) classical, (**S**) intermediate, and (**T**) non-classical monocytes (gated on total monocytes); (**U**) CD80, (**V**) CD163, and (**W**) CD206 expression on monocytes. Monocyte subset analysis was limited to 41 patients due to low monocyte frequencies. Each dot represents one patient. Data are shown as median with interquartile range. Statistical comparisons were performed using the Mann-Whitney U test. *, *p* < 0.05; **, *p* < 0.01.

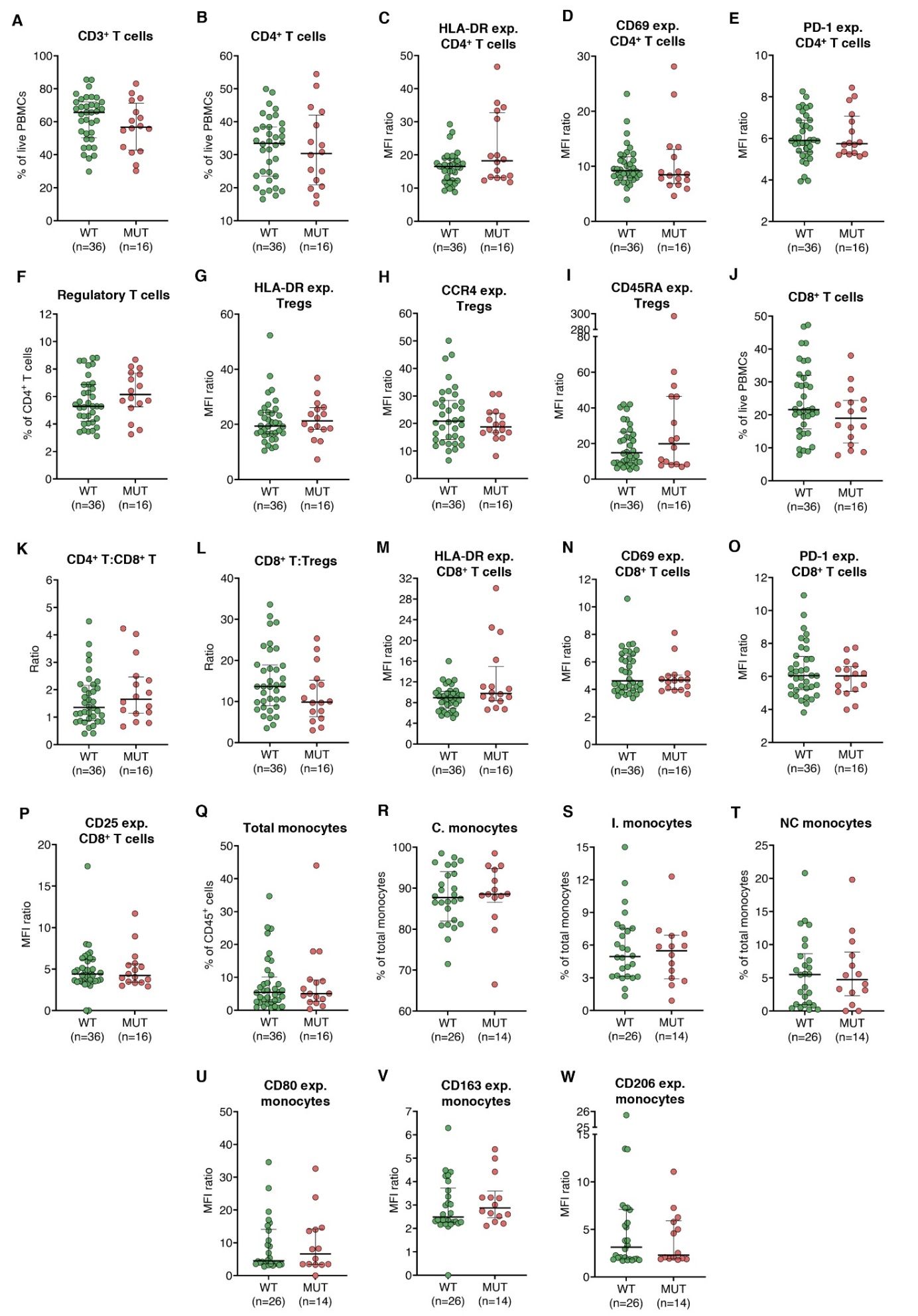
Supplementary Figure S5. Impact of *BRAF* mutational status on circulating immune cell subsets and functional marker expression in CM patients**.** Flow cytometry was used to quantify immune cell subsets and functional marker expression in PBMCs from 54 CM patients, stratified by *BRAF* mutational status. Marker expression levels (exp.) are defined as the ratio of MFI between marker-positive and -negative populations. Analyzed subsets and markers include (**A**) CD3^+^ T cells and (**B**) CD4^+^ T cells (gated on live PBMCs); (**C**) HLA-DR (**D**) CD69, and (**E**) PD1 expression on CD4^+^ T cells; (**F**) Tregs (gated on CD4^+^ T cells); (**G**) HLA-DR, (**H**) CCR4, and (**I**) CD45RA expression on Tregs; (**J**) CD8^+^ T cells (gated on live PBMCs); (**K**) CD4^+^:CD8^+^ T cell ratio and (**L**) CD8^+^ T cell:Treg ratio (gated on CD3^+^ T cells); (**M**) HLA-DR, (**N**) CD69, (**O**) PD-1, and (**P**) CD25 expression on CD8^+^ T cells; (**Q**) total monocytes (gated on CD45^+^ cells); (**R**) classical, (**S**) intermediate, and (**T**) non-classical monocytes (gated on total monocytes); (**U**) CD80, (**V**) CD163, and (**W**) CD206 expression on monocytes. Monocyte subset analysis was limited to 41 patients due to low monocyte frequencies. Each dot represents one patient. Data are shown as median with interquartile range. Statistical comparisons were performed using the Mann-Whitney U test.

Abbreviations: CM, cutaneous melanoma; MFI, median fluorescence intensity; MUT, mutated; PBMCs, peripheral blood mononuclear cells; Tregs, regulatory T cells; WT, wildtype.

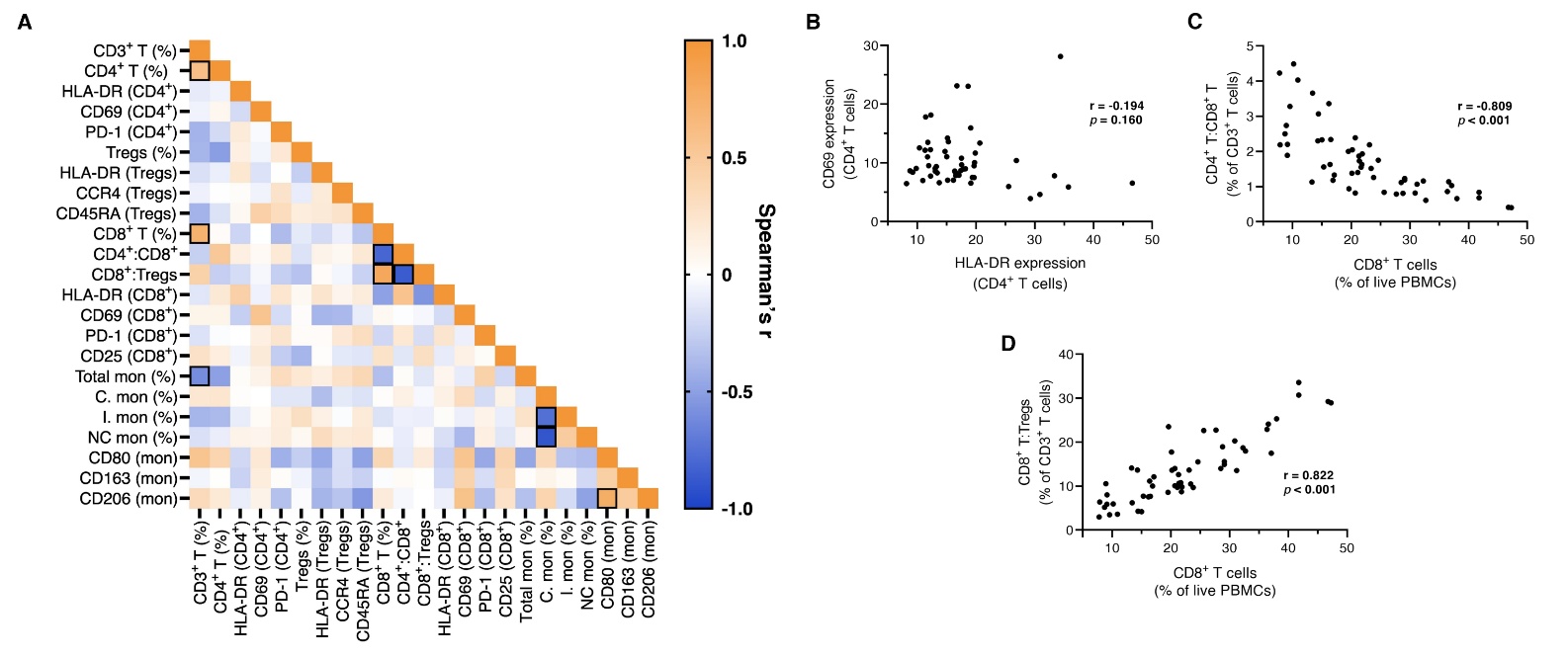
**Supplementary Figure S6. Correlation analysis identifies strongly associated circulating immune variables.** Spearman’s rank correlations were calculated across 54 treatment-naïve cutaneous melanoma patients, based on the frequency of major immune subsets and expression levels of functional markers, calculated by flow cytometry in PBMCs. Due to low monocyte frequencies in some individuals, monocyte subset analysis was limited to 41 CM patients. Expression levels are defined as the MFI ratio between marker‑positive and ‑negative populations. (**A**) Correlation matrix depicting Spearman’s rank correlation coefficients (r) for each pairwise comparison; significant correlations (*p* < 0.05 and |r| > 0.6) are outlined. (**B**) HLA-DR vs. CD69 expression on circulating CD4^+^ T cells. (**C**) Frequency of circulating CD8^+^ T cells (gated on live PBMCs) vs CD4^+^:CD8^+^ T cell ratio (gated on CD3^+^ T cells). (**D**) Frequency of circulating CD8^+^ T cells (gated on live PBMCs) vs. CD8^+^ T:Treg ratio (gated on CD3^+^ T cells).

Abbreviations: C., classical; I., intermediate; MFI, median fluorescence intensity; mon, monocytes; NC, non-classical; PBMCs, peripheral blood mononuclear cells; Tregs, regulatory T cells.

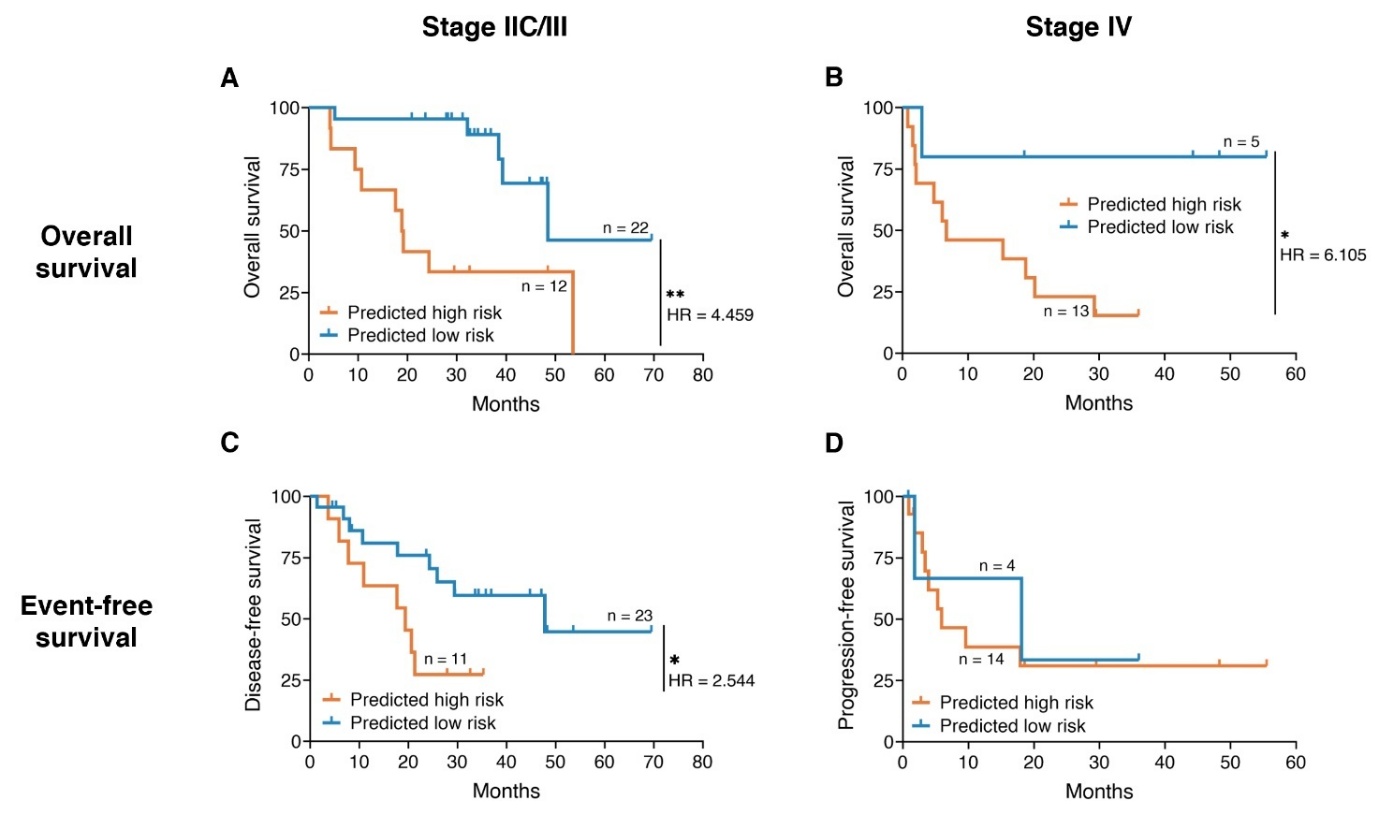
Supplementary Figure S7. Stage-specific risk stratification of cutaneous melanoma patients using final prognostic models. 52 treatment-naïve cutaneous melanoma patients were stratified by stage: 34 stage IIC/III patients (top panels) and 18 stage IV patients (bottom panels). Patients were divided into high- and low-risk groups using optimal cut-off values (maximized Youden’s index) derived from the final prognostic OS and EFS models. Kaplan-Meier survival curves with log-rank testing are shown for (**A,B**) OS, (**C**) DFS, and (**D**) PFS.

Abbreviations: DFS, disease-free survival; EFS, event-free survival; HR, hazard ratio; OS, overall survival; PFS, progression-free survival. *, *p* < 0.05; **, *p* < 0.01.

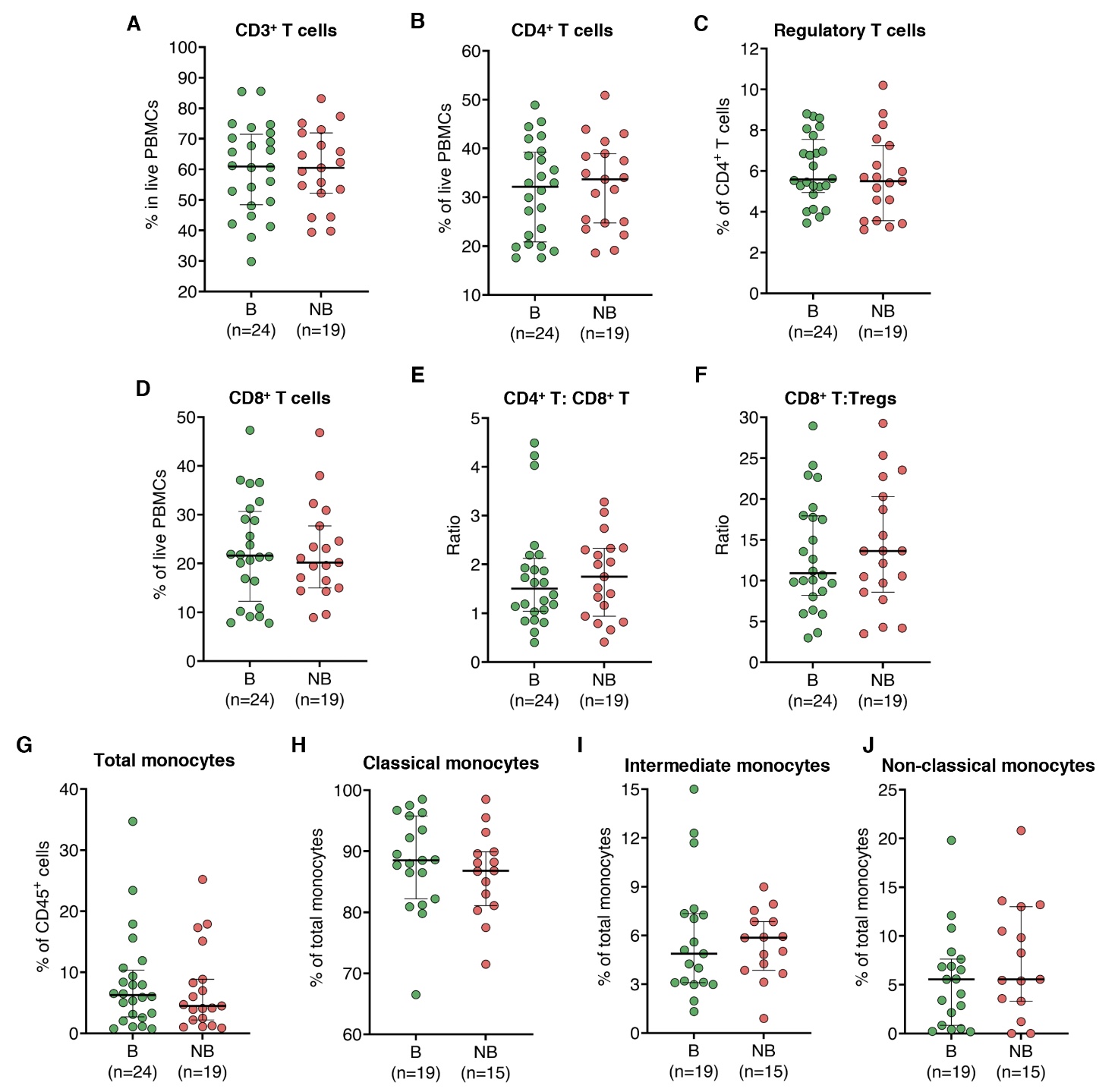
Supplementary Figure S8. Frequencies of circulating immune cell subsets at baseline do not differ between cutaneous melanoma patients with or without therapeutic benefit from ICIs**.** Percentages of immune cell subsets were quantified by flow cytometry in PBMCs from 43 CM patients prior to receiving ICI treatment, stratified by therapeutic benefit (yes vs. no). Analyzed subsets include (**A**) CD3^+^ T cells and (**B**) CD4^+^ T cells (gated on live PBMCs); (**C**) Tregs, gated on CD4^+^ T cells; (**D**) CD8^+^ T cells (gated on live PBMCs); (**E**) CD4^+^:CD8^+^ T cell ratio and (**F**) CD8^+^ T cell:Treg ratio (gated on CD3^+^ T cells); (**G**) total monocytes (gated on CD45^+^ cells); and (**H**) classical, (**I**) intermediate, and (**J**) non-classical monocyte subsets (gated on total monocytes). Each dot represents a patient. Data are shown as median with interquartile range. Statistical comparisons were performed using the Mann-Whitney U test.

Abbreviations: B, therapeutic benefit; CM, cutaneous melanoma; ICIs, immune-checkpoint inhibitors; NB, no therapeutic benefit; PBMCs, peripheral blood mononuclear cells; Tregs, regulatory T cells.

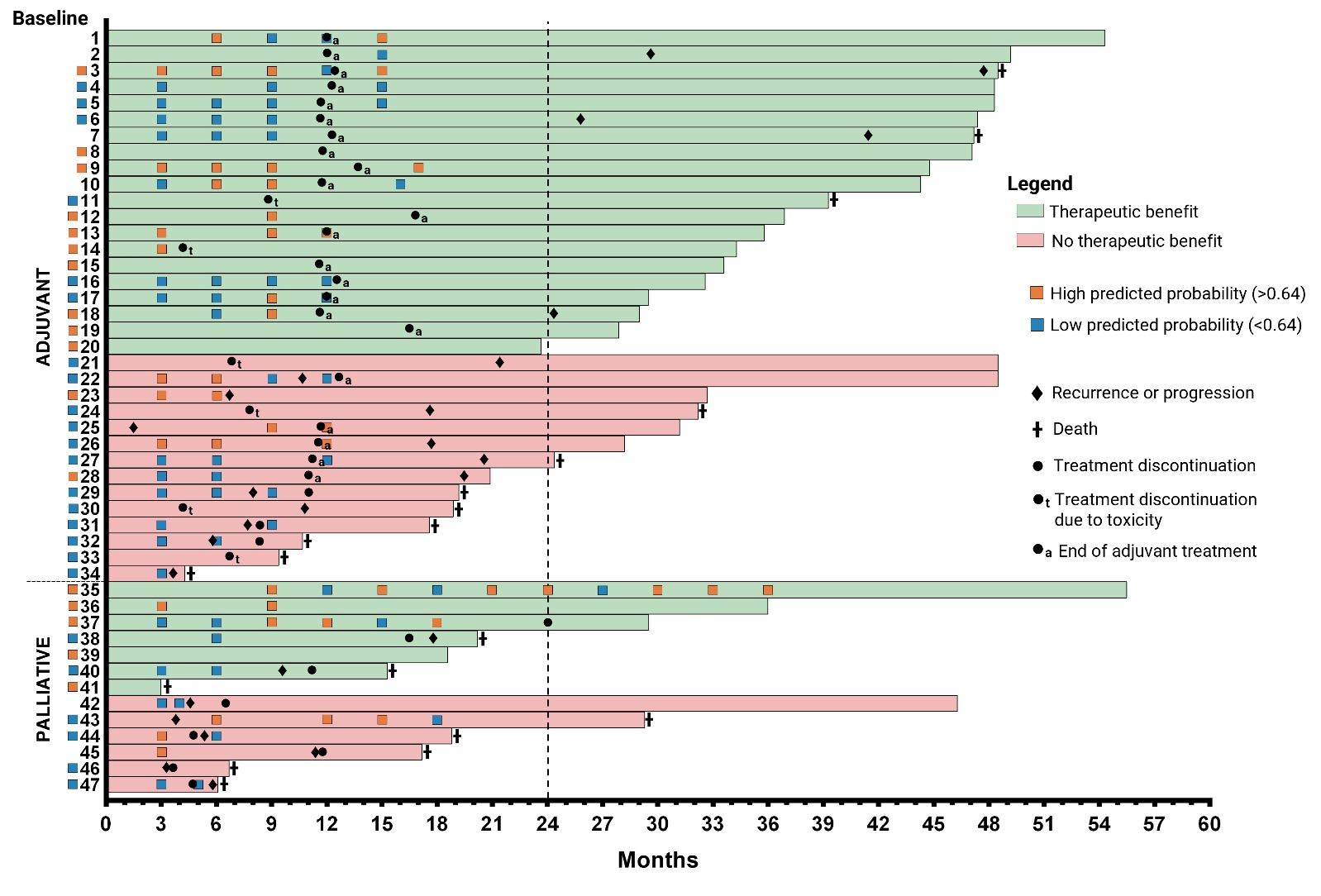
Supplementary Figure S9. Predictive model based on HLA-DR and CD69 expression on circulating CD4+ T cells lacks reliability for longitudinal monitoring during immune-checkpoint blockade**.** Swimmer plot depicting the evolution of predictive probabilities of therapeutic benefit during treatment in 47 cutaneous melanoma patients, as calculated by the previously established multivariable logistic regression model (two patients excluded due to missing clinical data). Probabilities were derived from HLA-DR and CD69 expression on CD4^+^ T cells (MFI ratio, flow cytometry) and the clinical risk score, and divided into high (orange squares) or low (blue squares) based on the optimal baseline cut-off (P = 0.64). Each horizontal bar represents one patient’s follow-up duration (in months) from the start of treatment, with the squares to the left of each bar indicating values at baseline. Key clinical events, such as progression, death, treatment discontinuation or termination, are annotated along the timeline.

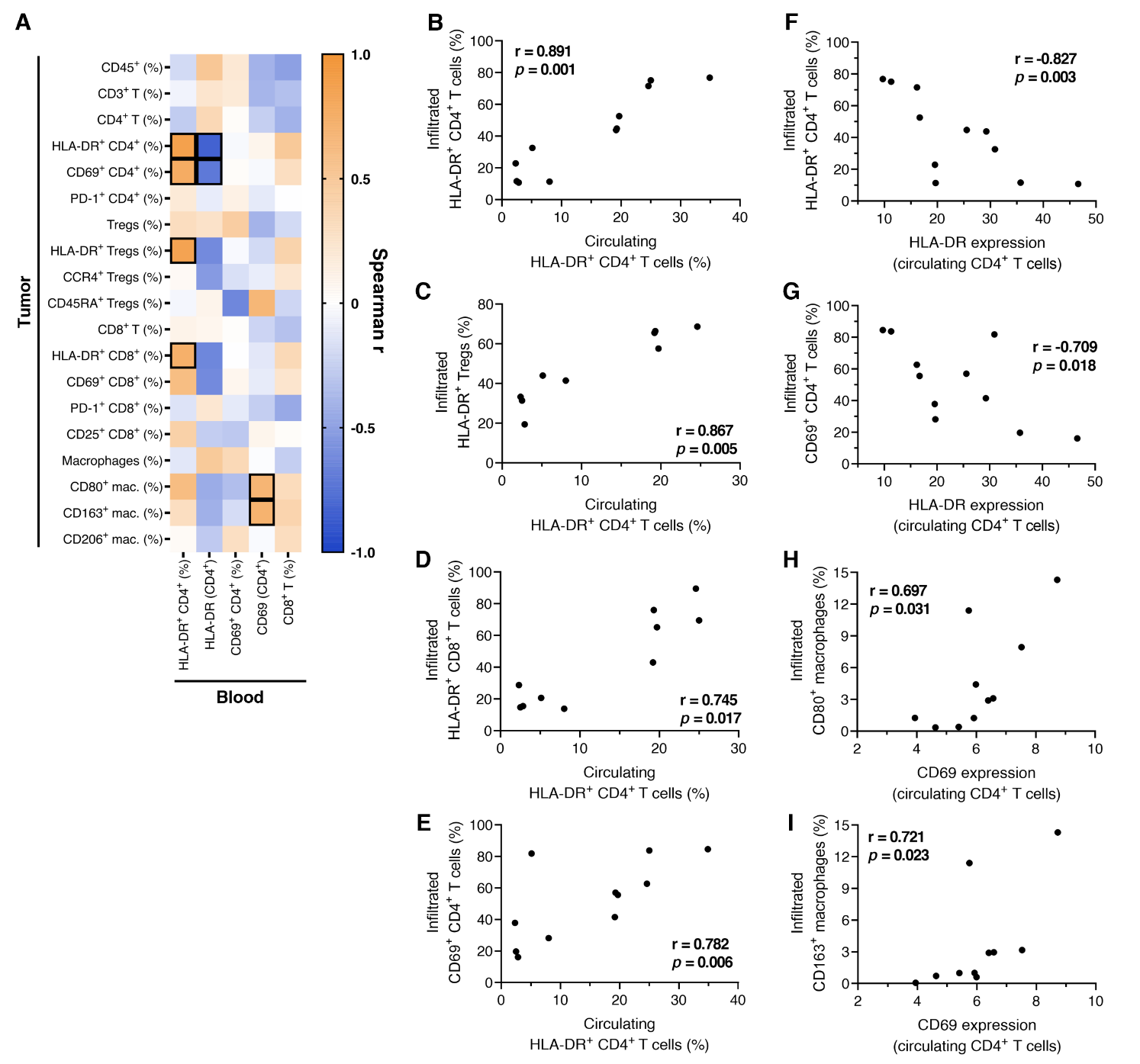

Supplementary Figure S10. The circulating immune profile partially reflects the tumor immune microenvironment**.** Spearman’s rank correlations were performed between circulating immune biomarkers of interest and the frequency of tumor‑infiltrating immune subsets, assessed by flow cytometry in paired blood and tumor samples from 11 cutaneous melanoma patients. Expression levels are defined as the median fluorescence intensity (MFI) ratio between marker‑positive and -negative populations. (**A**) Heatmap showing Spearman’s rank correlation coefficients (r) for each pairwise comparison; significant correlations (*p* < 0.05 and |r| > 0.6) are outlined. Correlations between the frequency of circulating HLA-DR^+^ CD4^+^ T cells (gated on CD4^+^ T cells) and the frequency of infiltrating (**B**) HLA‑DR^+^ CD4^+^ T cells, (**C**) HLA-DR^+^ Tregs, (**D**) HLA-DR^+^ CD8^+^ T cells, and (**E**) CD69^+^ CD4^+^ T cells (all gated on respective parent populations). Correlations between HLA-DR expression on circulating CD4^+^ T cells and (**F**) HLA-DR^+^ and (**G**) CD69^+^ tumor-infiltrating CD4^+^ T cells (gated on CD4^+^ T cells). Correlations between CD69 expression on circulating CD4^+^ T cells and (**H**) CD80^+^ and (**I**) CD163^+^ macrophages in the tumor (gated on CD68^+^ cells).

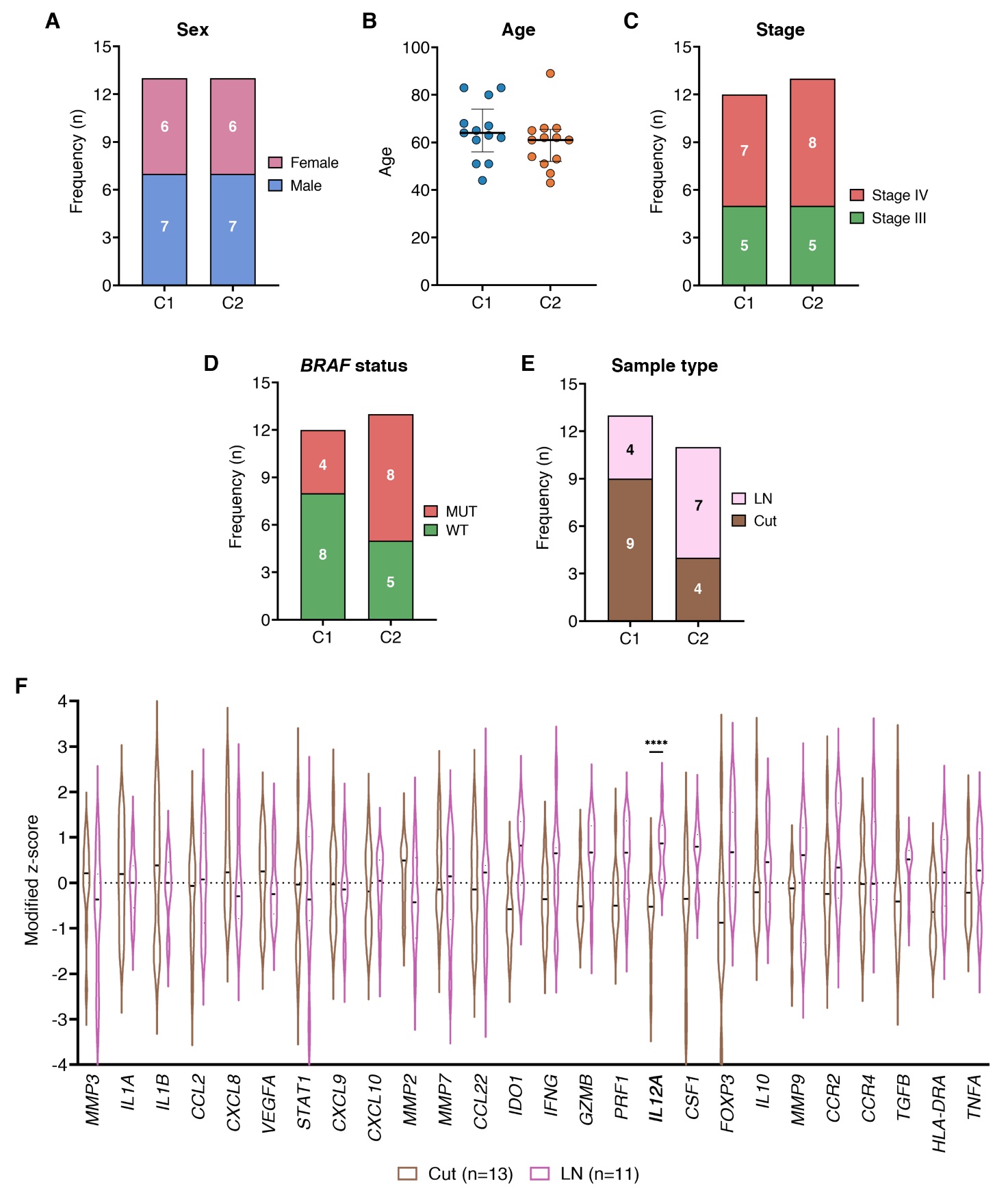
Supplementary Figure S11. Demographic and clinical characteristics of cutaneous melanoma patients according to tumor gene expression clusters**.** Clinical features of 26 cutaneous melanoma patients included in the tumor gene expression analysis, stratified by unsupervised clustering based on a 26-gene immune-related panel. (**A**) Distribution of sex between cluster 1 (C1) and 2 (C2). (**B**) Age distribution of patients in each cluster, presented as median with interquartile range. (**C**) Distribution of disease stages in each cluster; 1 patient with stage IIC excluded. (**D**) Frequency of *BRAF* mutational status (WT, wildtype; MUT, mutant) in each cluster; 1 patient excluded due to missing data. (**E**) Distributions of lymph node (LN) and cutaneous (Cut) samples in each cluster; 2 patients with soft tissue metastases excluded. (**F**) Violin plots showing the distribution of modified z-scores for individual immune-related genes in cutaneous versus lymph node samples. Associations between cluster assignment and binary clinical variables were assessed using Fisher’s exact test; age comparisons between clusters were performed using the Mann-Whitney U test; differences in gene expression between sample types were assessed by multiple Mann-Whitney U tests, followed by false discovery rate correction using the Benjamini‑Kieger‑Yekutieli two-stage step-up method. ****, *p* < 0.0001.

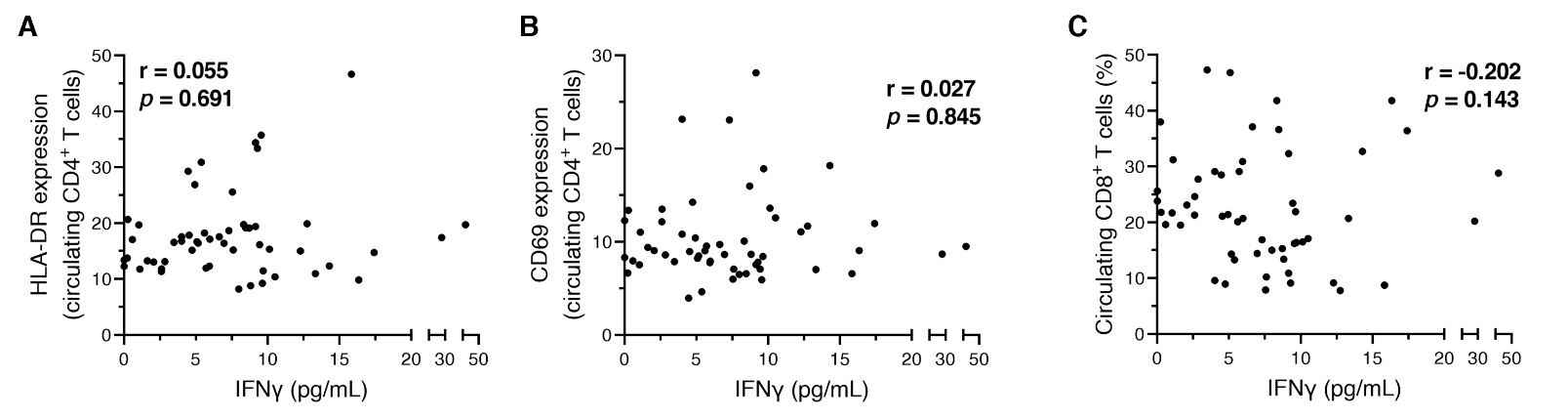
Supplementary Figure S12. Correlation between identified circulating prognostic biomarkers and plasma IFN-γ levels**.** Spearman’s rank correlations were performed using samples from 54 treatment-naïve cutaneous melanoma patients. Circulating immune biomarkers of interest were measured in peripheral blood mononuclear cells (PBMCs) by flow cytometry, while plasma IFN-γ concentrations were quantified by ELISA. Biomarker expression levels are defined as the median fluorescence intensity (MFI) ratio between marker‑positive and -negative populations. Correlations were assessed between plasma IFN-γ concentrations and (**A**) HLA-DR expression on circulating CD4^+^ T cells, (**B**) CD69 expression on circulating CD4^+^ T cells, and (**C**) frequency of circulating CD8^+^ T cells (gated on live PBMCs). Each dot represents a patient.
